# Deeplasia: prior-free deep learning for pediatric bone age assessment robust to skeletal dysplasias

**DOI:** 10.1101/2023.03.07.23286906

**Authors:** Sebastian Rassmann, Alexandra Keller, Kyra Skaf, Alexander Hustinx, Ruth Gausche, Miguel A. Ibarra-Arrelano, Tzung-Chien Hsieh, Yolande E. D. Madajieu, Markus M. Nöthen, Roland Pfäffle, Ulrike I. Attenberger, Mark Born, Klaus Mohnike, Peter M. Krawitz, Behnam Javanmardi

## Abstract

**Background:** Skeletal dysplasias collectively affect a large number of patients worldwide. The majority of these disorders cause growth anomalies. Hence, assessing skeletal maturity via determining the bone age (BA) is one of the most valuable tools for their diagnoses. Moreover, consecutive BA assessments are crucial for monitoring the pediatric growth of patients with such disorders, especially for timing hormone treatments or orthopedic interventions. However, manual BA assessment is time-consuming and suffers from high intra-and inter-rater variability. This is further exacerbated by genetic disorders causing severe skeletal malformations. While numerous approaches to automatize BA assessment were proposed, few were validated for BA assessment on children with abnormal development.

**Objective:** We design and present Deeplasia, an open-source prior-free deep-learning approach for pediatric bone age assessment specifically validated on patients with skeletal dysplasias.

**Materials and methods:** We extensively experiment with training multiple convolutional neural network models under various conditions and select three to build a precise model ensemble. We utilize the public RSNA BA dataset consisting of training, validation, and test subsets each containing 12,611, 1,425, and 200 hand X-rays, respectively. For testing the performance of our model ensemble on dysplastic hands, we retrospectively collected 568 X-ray images from 189 patients with molecularly confirmed diagnoses of seven different genetic bone disorders including Achondroplasia and Hypochondroplasia.

**Results:** On the public RSNA test set, we achieve state-of-the-art performance with a mean absolute difference (MAD) of 3.87 months based on the average of six different reference ratings. We demonstrate the generalizability of Deeplasia to the dysplastic hands (unseen by the models) achieving a MAD of 5.84 months w.r.t. to the average of two reference ratings. Further, using longitudinal data from a subset of the dysplastic cohort (149 images), we estimate the test-retest precision of our model ensemble to be at least at the human expert level (2.74 months).

**Conclusion:** We conclude that Deeplasia suits assessing and monitoring the BA in patients with skeletal dysplasia.

## 1 Introduction

The estimation of bone age (BA), which evaluates skeletal maturity, is one of the most important and valuable tools in assessing children’s health. Usually, it is one of the first steps in the diagnosis of pediatric growth disorders (Creo and Schwenk, 2017). In particular, for conditions in which hormonal therapy or orthopedic interventions are being considered, the timing of the treatment depends on the assessed BA (Bunch et al, 2017). The BA can be estimated by observing the ossification centers of a child’s skeleton. The main body parts used for BA assessment are the hands, wrists, and knees. BA estimates from the hand and wrist are more closely correlated with the child’s overall growth progress and puberty onset than estimates from the knee. Hence, the BA estimated from hand X-rays is more effective in assessing delayed or advanced growth (Aicardi et al, 2000) and is therefore used as a routine diagnostic and monitoring method (Martin et al, 2011a,b). The Greulich-Pyle (hereafter GP, Greulich and Pyle, 1959) and Tanner-Whitehouse (hereafter TW, Tanner, 1962; Tanner et al, 1975, 2001) are the two most commonly used hand and wrist BA estimation methods. While the TW method is considered to be more accurate, the GP method is generally regarded to be faster (De Sanctis et al, 2014). Nevertheless, both methods are time-consuming and show high degrees of inter- and intra-rater variability (De Sanctis et al, 2014; Eng et al, 2021).

Artificial Intelligence (AI) methods are contributing to all medical fields (Rajpurkar et al, 2022) including pediatric radiology (Offiah, 2021) and numerous Machine Learning (ML) approaches were proposed to automate BA assessment, especially based on a publicly available dataset released in 2017 by the Radiological Society of North America (RSNA) for their pediatric BA challenge (Larson et al, 2018; Halabi et al, 2019). While an approach using end-to-end Deep Learning (DL) without any human prior or a particularly task-specific design won the competition (Halabi et al, 2019; Cicero and Bilbily, 2017), ML approaches emphasizing anatomical features used in human BA assessment showed some improvement in more recent studies (e.g. Escobar et al, 2019; Wang et al, 2020a; Koitka et al, 2020; Martin et al, 2022).

A major indication to perform BA assessments are suspected growth or developmental anomalies. This is often connected with the phenotype of a skeletal dysplasia (Spranger et al, 2018) due to the common causation by a genetic disorder. Although these disorders are individually rare (Unger et al, 2023), collectively they affect a large number of children (Sabir and Cole, 2019) with an estimated total number of around 25 million worldwide. Especially in such patients, reliable and precise BA estimations are vital, both, to obtain an initial diagnosis and to monitor the maturation progress over time (Satoh and Hasegawa, 2022). As skeletal dysplasias obstruct precise BA assessment (Martin et al, 2011a), automated and ML-based tools require specific validation on patients with such disorders. However, this problem is still understudied and many approaches to automatic BA assessment were developed for and tested on datasets composed of predominantly normally-developing children. The public dataset released as part of the 2017 BA challenge contains only 0.21% cases of reported skeletal dysplasias (Larson et al, 2018; Halabi et al, 2019) and the more recent study by Eng et al (2021) included *<* 1.4% patients with congenital diseases. Kim et al (2017) and Wang et al (2020b) proposed and tested DL methods on patients with abnormal growth, however, their study was limited to Korean and Chinese populations, respectively, and the test sets included no or only small numbers (*n <* 10) of images from patients with severe skeletal malformations such as Achondroplasia (ACh) and Hypochondroplasia (HyCh). Furthermore, the altered hand morphology impedes the automatic identification of individual bones or regions of interest (ROIs) in dysplastic hands. Hence, ML approaches relying on the identification of particular ROIs within the hand might be unsuitable for precise BA assessment on dysplastic hands. For example, the routinely-used BA assessment tool BoneXpert (Visiana, Hørsholm, Denmark, Thodberg et al, 2009) rejects some of the malformed hands and struggles to generalize to all patients with skeletal dysplasias such as ACh (Offiah and Hall, 2020; Offiah, 2021).

In this work, we design and introduce Deeplasia: an AI application for pediatric BA assessment specifically validated on dysmorphic hands of patients with skeletal dysplasias. Given the intrinsic scarcity of data from patients with rare diseases, we aim at presenting an open-source tool that while trained on data of normal hands can reliably be used for assessing BA of patients with rare bone diseases. To this end, and to overcome the dependence on potentially corrupted individual features commonly used for human BA assessment, we employ an end-to-end DL approach. This allows learning a broader range of features rather than emphasizing or completely depending on predefined shapes or ROIs. We demonstrate that our prior-free learning approach is at least as powerful as other approaches which incorporate priors from human BA assessment and which require extensive additional annotation.

We show that Deeplasia

- achieves state-of-the-art (SOTA) performance on the RSNA BA test dataset composed of predominantly healthy patients,
- generalizes to patients from unseen cohorts and with a variety of genetically-confirmed skeletal dysplasias,
- is demonstrated to be applicable to longitudinal data from patients with skeletal dysplasias for the purpose of progressive growth monitoring, and
- can assess the BA in less than 10 seconds on consumer-grade hardware without GPU acceleration.

## 2 Materials and methods

### 2.1 Training and validation datasets

We use the 2017 RSNA training and validation sets containing 12,611 and 1,425 images, respectively. RSNA published these data for their Pediatric BA ML Challenge (Larson et al, 2018; Halabi et al, 2019). The data were obtained from Children’s Hospital Colorado (Aurora, CO, USA) and Lucile Packard Children’s Hospital at Stanford (Palo Alto, CA, USA). For each image, the sex and a ground truth GP BA estimate are provided. For determining the ground truth BA, one estimate from the original clinic of the data, a second estimate from the same rater at least one year later, and four independent estimates were obtained. To form the final consensus BA estimate, a weighted mean based on the performance of each reviewer is calculated (for more details see Halabi et al, 2019). The mean chronological age of patients in the training and validation set is 10.8 *±* 3.5 years (Larson et al, 2018) and their mean estimated BA is 10.6 *±* 3.4 years (Halabi et al, 2019).

### 2.2 Test datasets

For validating our AI, we used three independent test datasets as described below.

#### 2.2.1 RSNA test set

The RSNA test dataset from the Pediatric BA ML Challenge (Larson et al, 2018; Halabi et al, 2019) contains 200 images (100 males and 100 females) from Lucile Packard Children’s Hospital. The mean chronological age of patients in this set is 11.3 *±* 3.8 years (Larson et al, 2018), and the mean estimated BA is 11.0 *±* 3.6 years (Halabi et al, 2019). Similar to the training and validation sets, the sex and a ground truth GP BA estimate (weighted average of six measurements) are provided for each image. The distribution of ground truth BA for males and females in the test set is similar to those in the training and validation sets.

#### 2.2.2 Los Angeles Digital Hand Atlas

As an additional test set for normally-developing children, we used the publicly released Los Angeles Digital Hand Atlas (DHA, Gertych et al, 2007; Zhang et al, 2009). It consists of 1,390 images acquired between 1997 and 2008 at the Children’s Hospital Los Angeles, USA. The study cohort included four ethnicities and ground truth BA estimates were obtained by two raters using the GP atlas. The ground truth BA was defined as the average of the two ratings. We excluded seven images due to lacking or completely implausible ground truth BA assessment (BA of 99 years, BA of 0 years for children of 9 years of chronological age, and two images with a difference to a third manual assessment by K.M. of *>* 2 years).

#### 2.2.3 German Dysplastic Bone Dataset

To compile a dataset for validating the BA prediction models on dysplastic hands, we retrospectively (2006 -2022) collected hand X-rays from patients referred to the pediatric endocrinology of two German university hospitals (Magdeburg and Leipzig) due to a suspected growth disorder. The X-ray images were acquired as hard copies and thereafter digitized. The study was approved by the ethics committee of the medical faculties of the universities Magdeburg (vote 27/22) and Leipzig (vote 121/22-ek).

We term this dataset the German Dysplastic Bone Dataset (GDBD). In total, it contains 568 hand X-ray images from 189 patients with molecularly confirmed diagnoses of one of the following disorders: ACh, HyCh, Pseudohypoparathyroidism (PsHPT), Noonan, Silver-Russel (SRS), and Ullrich-Turner (UTS) syndromes, and a mutation in the SHOX gene. Further, 55 images from 12 patients with intrauterine growth restriction (IUGR) were included. The number of images and patients as well as the distribution of their chronological age are shown in Fig. 1. An example X-ray per each disorder is shown in Fig. 2. We also included 79 images from 79 children without a diagnosed growth disorder, but who had been referred to pediatric endocrinologists due to a suspected growth disorder. The ethnic background of the GDBD patients is not available, however, we suspect a large portion of them to be Caucasians.

**Fig. 1.**
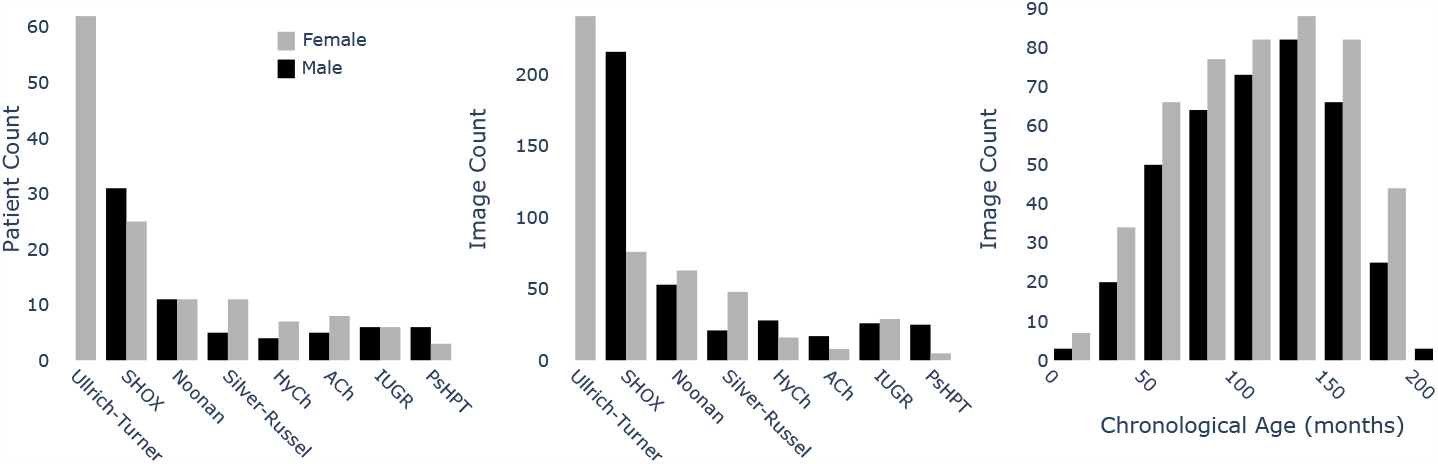
The GDBD distribution of patient and image counts between disorders, and the cross-cohort distribution of chronological age. ACh: Achondroplasia, HyCh: Hypochondroplasia, PsHPT: Pseudohypoparathyroidism, and IUGR: intrauterine growth restriction.

**Fig. 2.**
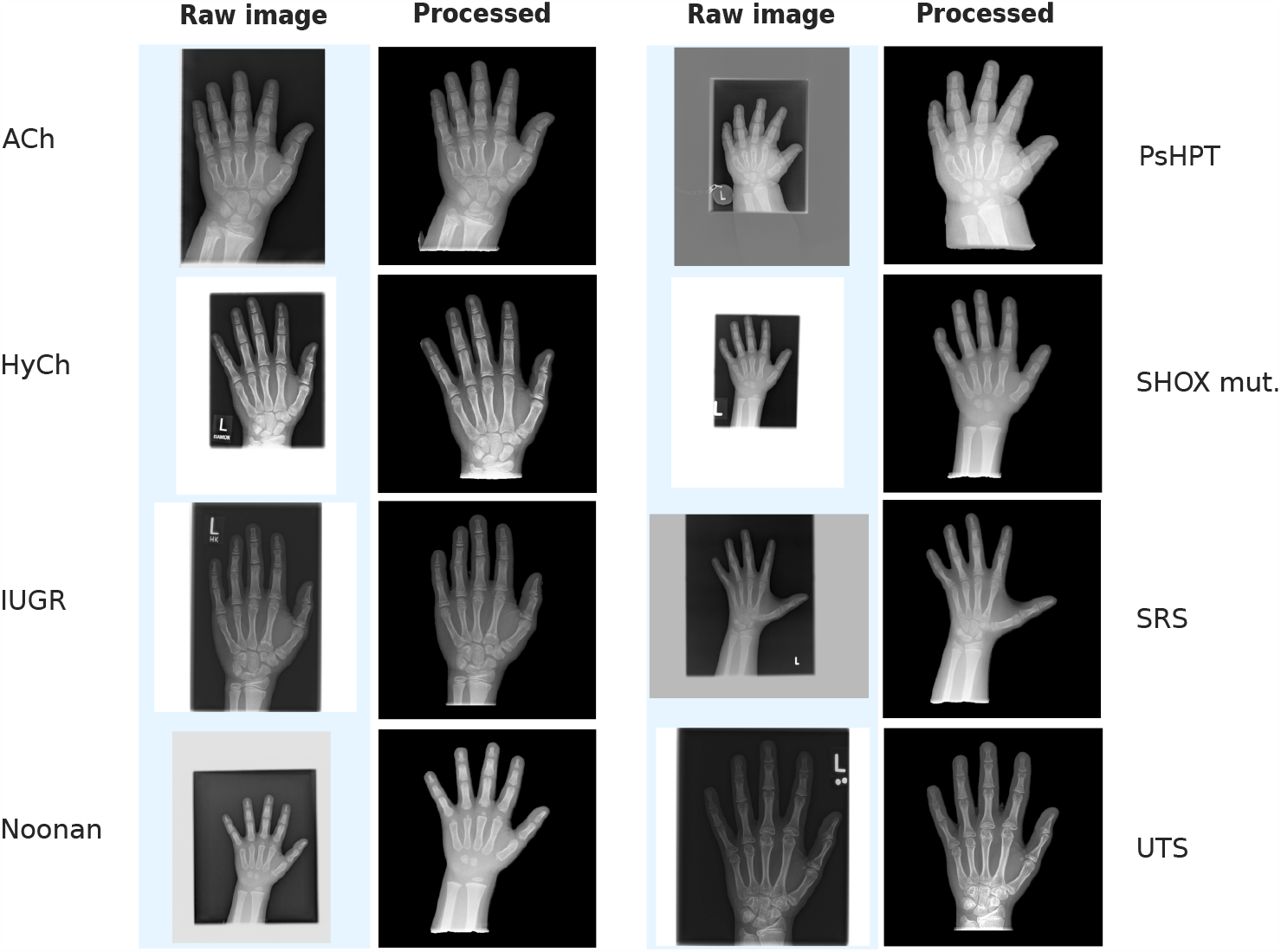
Example hand X-ray for each bone dysplasia in the GDBD. For each image, the original raw image and the preprocessed version (see Section 2.3.1) are shown. Note, that the hands have a wide range of relative scales within the images, image qualities vary, and show artifacts such as labels and white or gray backgrounds caused by scanning. ACh: Achondroplasia, HyCh: Hypochondroplasia, PsHPT: Pseudohypoparathyroidism, IUGR: intrauterine growth restriction, SHOX mut.: mutation of the SHOX gene, SRS: Silver-Russel Syndrome, and UTS: Ullrich-Turner Syndrome.

The BA reference grading for the GDBD was obtained using the GP standard by two pediatric endocrinologists with more than 40 (K.M.) and more than 20 (A.K.) years of clinical experience. For 643 out of 702 images, one of the assessments was obtained from the initial clinical report. The BA ratings for the remaining images and the second reference ratings were obtained from a dedicated session, in which the images were presented a) using the same preprocessing procedures as for testing the models (see Section 2.3.1), b) in a randomized order, and c) blinded for the chronological age, the clinical report, and the diagnosis.

Figure 3 shows how the above-described datasets were used in the training and testing of our AI.

**Fig. 3.**
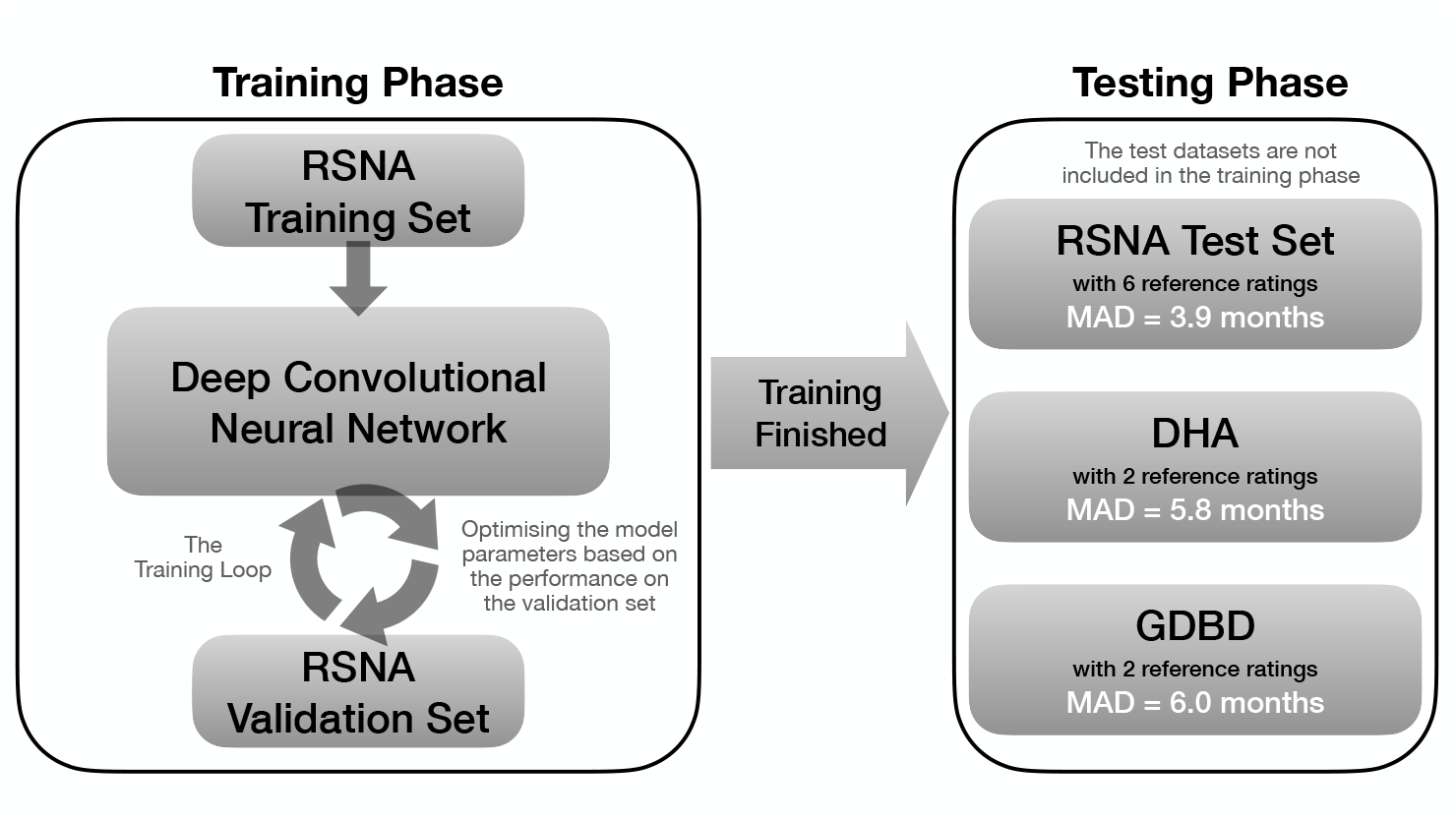
A flowchart describing the use of the different datasets described in Section 2 for training and testing of our models. MAD: mean absolute difference (see the Results section).

### 2.3 Design and development of Deeplasia

#### 2.3.1 Image background removal and preprocessing

Imaging and scanning induce artifacts to the input images, e.g. department-specific markers or white borders surrounding the X-ray carrier in the scan. Such artifacts (which are present in many of the images in our GDBD set) have been shown to bias DL models (e.g. Zech et al, 2018). Further, high-intensity borders can potentially skew the image normalization for inference. To prevent these problems, we trained and incorporated DL modules within Deeplasia to automatically extract the hand from the scan by masking the background. Fig. 2 shows some examples of the results of our preprocessing on dysplastic hands. It can be seen how the preprocessing removed the backgrounds and cropped the GDBD images to the predicted mask (to assert a constant scale of the hand in the follow-up analysis). The details of our hand segmentation approach are described in the supplementary information. The training masks and the code for the hand segmentation are publicly available via Rassmann et al (2023) and github.com/aimi-bonn/hand-segmentation, respectively.

#### 2.3.2 Bone age model training

As a baseline approach for the BA model, we follow the design principle winning the 2017 RSNA Pediatric BA ML Challenge (Halabi et al, 2019; Cicero and Bilbily, 2017). The model architecture, outlined in Fig. 1 of the supplementary information, is composed of a fully Convolutional Neural Network (CNN) as a feature extractor, channel-wise average pooling of the extracted features, and concatenation of a representation of the patient’s sex inflated to 32 neurons^1^. The results are passed through a variable set of fully-connected (FC) layers to achieve a final prediction. We employ *EfficientNets* (Tan and Le, 2019) as backbone feature extractors. In comparison to previously proposed end-to-end learning methods (Cicero and Bilbily, 2017; Torres et al, 2020), our applied average pooling reduces the dimensionality of the learned features and, thus, decreases the model size. For example, the largest of our BA models has a feature dimensionality of 1, 792 resulting in a total network size of 23 *×* 10^6^ parameters, while the configuration proposed by Torres et al (2020) uses a feature dimensionality of 33, 712 and 82 *×* 10^6^ parameters. All the details of our training procedure for reproducing the models are described in the supplementary information, and the open-source code for training the BA models is available at github.com/aimi-bonn/Deeplasia and deeplasia.de.

#### 2.3.3 Model explorations for building an ensemble

To build the model ensemble for Deeplasia, we experimented with three different training conditions, a) the *baseline*: *EfficientNet-b0* with 512 *×* 512 input resolution, b) *large CNN* : *EfficientNet-b4* with 512 *×* 512 input resolution, and c) *high-resolution*: *EfficientNet-b0* with 1024 *×* 1024 input. For each of these conditions, we trained models with three sets of FC layers: [256], [512, 512], and [1024, 1024, 512, 512]. Therefore, in total, we trained nine CNN models for BA estimation. Fig. 4 shows a flowchart describing this procedure and the details of our training experimentations are described in the supplementary information.

**Fig. 4.**
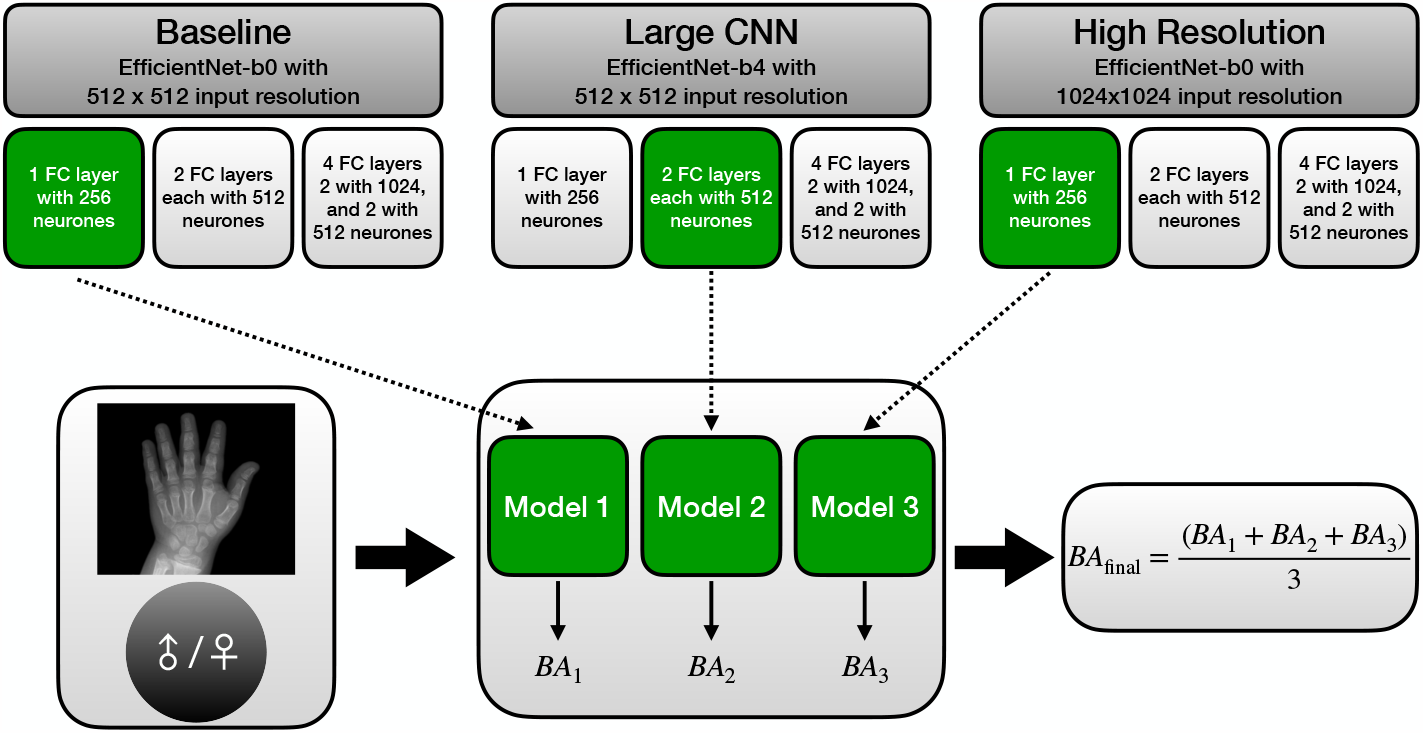
A flowchart describing the procedure for experimenting with different training configurations for building a model ensemble. We experimented with three different training conditions (*baseline, large CNN*, and *high-resolution*) each with three sets of FC-layer combinations. Out of these nine CNN models, we selected three (one per training condition, shown in green in the figure) based on their performance on the validation data set. See Section 2.3.3 and the supplementary information for further details. FC: fully connected, BA: bone age, CNN: convolutional neural network.

To choose the models for building an ensemble, we analyzed the pairwise correlations between the predicted BA of these nine models (visualized in Fig. 2 of the supplementary information). This revealed that there is a higher correlation between the predictions of the models within each training condition (i.e. the *baseline, large CNN*, and *high-resolution*) compared to the ones between the models across these conditions. As dissimilar prediction patterns in a model ensemble are advantageous due to partial compensation of predictive errors, it is beneficial to construct an ensemble composed of models across different training conditions. Consequently, we chose to pick the best-performing model from each of the three training conditions (shown by green boxes in Fig. 4) for building our model ensemble. The final BA is the average of the results from these three models.

### 2.4 Evaluation methods

#### 2.4.1 Metrics and statistical analysis

For model selection and benchmarking the mean absolute difference (MAD) was used. It is calculated as the *L*_1_-norm of the difference between predicted BA *Ŷ* = (*ŷ*_1_, *ŷ*_2_, …, *ŷ*_*n*_)^*T*^ and the respective ground truth *Y* = (*y*_1_, *y*_2_, …, *y*_*n*_)^*T*^ :

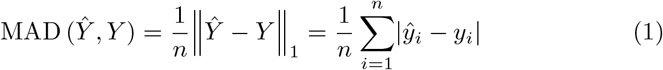

Further, the root-mean-square error (RMSE) was used as a metric that is more sensitive to outliers. It is defined as:

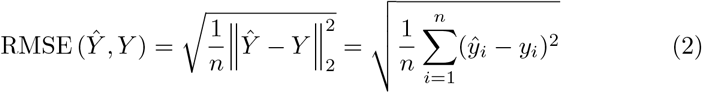

For the statistical analysis, we assume the signed error to be normally distributed and, thus, derive the confidence intervals of the RMSE from the corresponding *χ*^2^ distribution. As an additional, clinically more interpretable metric, we define a 1-year accuracy. Let **1**_*cond*_ denote the indicator function (a function that evaluates to 1 if and only if *cond* is true) and assume the BAs *Ŷ* and *Y* to be denoted in years, then

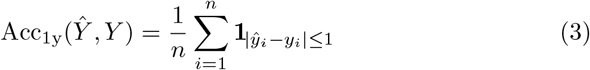

Note that we do not conduct a symbolic perturbation, so the measure is conservative w.r.t. the model performance as the models are, in contrast to human raters, unlikely to assign integer BAs.

#### 2.4.2 Longitudinal analysis

To detect small changes in the developmental process like slow-downs or growth spurts, BA measurements are required to have high test-retest reliability. Directly measuring the test-retest reliability would require a dedicated imaging session which would be unethical due to the unnecessary radiation exposure. However, assuming linear progress of the BA over time, the test-retest reliability can be estimated retrospectively from regular check-ups within the testing cohort. For estimating the upper bound of the expected error in assessing the BA, the method proposed by Thodberg and Sävendahl (2010) was used. No patients were excluded due to therapies or other interventions. Additionally, the potentially variable growth patterns due to the disorders of the patients included in the analysis might give a non-linear growth pattern. To account for this, we set the maximum time difference for the derivation triplets to 14 months, the lowest threshold to achieve *n ≥* 100 triplets. For analyzing the rater performance, only triplets derived from either the clinical ratings or from a single rater within the blinded re-rating session were included to avoid rater-rater biases or biases between clinical and blinded reviews.

## 3 Results

### 3.1 Performance on the RSNA test set

On the RSNA test set comprising 200 X-rays, Deeplasia achieved a MAD of 3.87 months, RMSE of 5.14 months, and a 1-year accuracy of 98.5%. These results, listed in Table 1, are on-a-par with the current SOTA (3.91 months, Wang et al, 2020a) and tools cleared for clinical use (4.1 months, Thodberg et al, 2009; Martin et al, 2022). Interestingly, even the individual three models of our ensemble (see Table 2 of the supplementary information) achieved test accuracies (MAD of 4.2, 4.1, and 4.3 months) comparable to other approaches incorporating human priors.

**Table 1.**
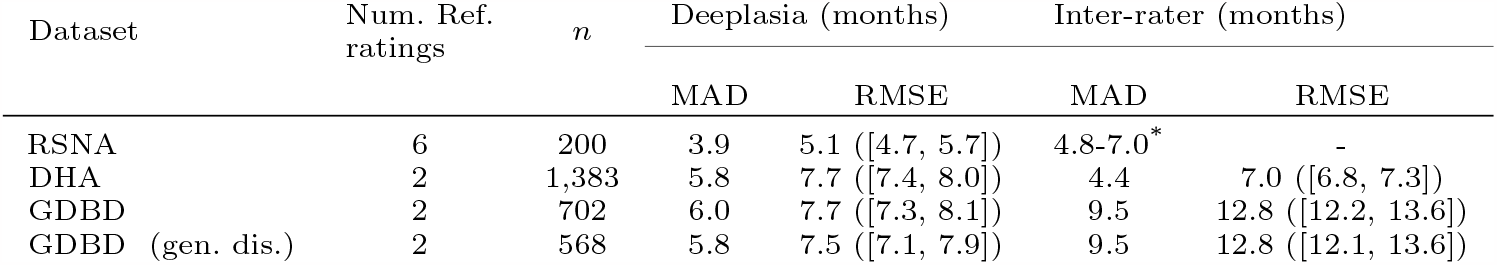
Deeplasia and inter-rater accuracies across different test datasets (lower MAD and RMSE errors mean higher accuracy). For the GDBD additionally, the scores for images from patients with molecularly confirmed genetic disorders (gen. dis.) are provided. The RMSE is stated with the 95% confidence interval. *n* refers to the number of individual X-rays in each set. ^*^Estimated range for the accuracies of the assessed single raters.

**Table 2.** The test-retest precision of Deeplasia, the clinical, and a blinded manual bone age rating is estimated on patients with genetically-confirmed disorders (i.e. excluding IUGR) within the GDBD. *n* refers to the number of images within the GDBD for which the interpolation residuals could be estimated. The precision is stated with the 95% confidence interval.

| Rating method | Precision (months) |  |
| --- | --- | --- |
| | full dataset ($n = 149$) | with clinical BA ($n = 106$) |
| Deeplasia | 2.7 ([2.5, 3.1]) | 2.4 ([2.1, 2.8]) |
| Clinical rating | - | 2.6 ([2.3, 3.0]) |
| Blinded rating | 5.6 ([4.9, 6.2]) | 5.8 ([5.1, 6.7]) |

### 3.2 Performance on the DHA dataset

To assess the generalizability to external test cohorts and potentially unseen ethnicities, we evaluated Deeplasia on the DHA dataset (Gertych et al, 2007; Zhang et al, 2009). We used 1,383 X-ray images from children (age 0-18 years) with different ethnic backgrounds and their corresponding BA ratings. On this dataset, Deeplasia achieves a MAD of 5.81 months, RMSE of 7.67 months, and a 1-year accuracy of 92.9% (see the second row of Table 1). Note that for this dataset the ground truth BA estimates are based on two rather than six raters for the RSNA test set.

### 3.3 Performance on the GDBD

Finally, we evaluated the performance on the GDBD to assess the generalization of Deeplasia to patients with skeletal malformations. Overall, the GDBD contains 568 images from patients with a molecularly confirmed genetic disorder, 55 images from patients with IUGR, and 79 images from individuals without any known disorders, but who had been referred to pediatric endocrinologists due to a suspected growth disorder. All reference BA ratings were performed by the same two raters (K.M. and A.K.).

Comparing the predictions of Deeplasia and the ground truth estimates defined by the average of two raters, the model ensemble achieves a MAD of 5.96 months, RMSE of 7.67 months, and a 1-year accuracy of 90.2% for the full set and 5.84 months (MAD), 7.48 months (RMSE), and 90.1% (1-year accuracy) for the subset of patients with molecularly confirmed disorders. These values (also listed in the third and fourth rows of Table 1) are similar to those from the performance on the DHA dataset and in the range of the single rater estimated in the annotation of the RSNA BA challenge (Halabi et al, 2019). Consequently, the error of the model ensemble w.r.t. the average of two reference ratings is smaller than the assessed inter-rater error (Table 1). In the left panel of Fig. 5, we illustrate the *Bland-Altman* plot for Deeplasia. It shows the difference between the BA predictions from Deeplasia and the reference values (from the two raters) vs. the average of the two methods. The signed mean difference of the two methods is 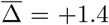 months (shown by a dotted line), and the plot reveals no systematic over- or underestimation of the BAs for different skeletal disorders. The difference between the predicted BA and the reference ratings is within 1.96 standard deviations (i.e. the 95% confidence interval) for 95.6% of the predicted BAs.

**Fig. 5.**
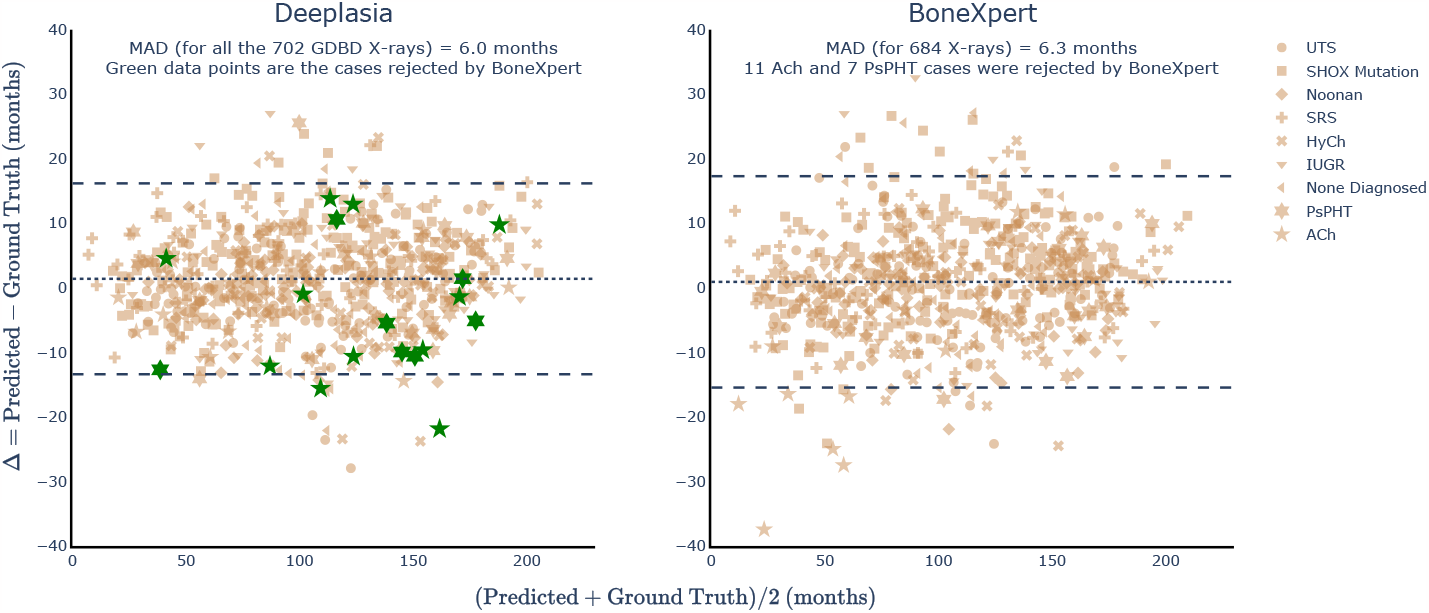
*Bland-Altman* plots showing the performances of Deeplasia (left) and BoneXpert (right) on the GDBD hand X-rays. On each panel, the Y axis is the difference between the software predictions and the ground truth values, and the X axis is their average. The dotted lines denote the mean (signed) difference (1.4 and 0.9 months for Deeplasia and BoneXpert, respectively). The dashed lines show the 95% or 1.96*σ* confidence intervals. Individual disorder categories are shown by different symbols as indicated in the legend. The green data points on the left panel show the 11 Ach and 7 PsHPT cases that were rejected by BoneXpert. For these cases, the performance of Deeplasia is similar to that for other cases and 16 out of 18 of them are within the 95% confidence region. On the other hand, even for the Ach cases that were not rejected by BoneXpert, a drop in its performance is visible for children below 5 years old. ACh: Achondroplasia, HyCh: Hypochondroplasia, PsHPT: Pseudohypoparathyroidism, IUGR: intrauterine growth restriction, SHOX mut.: mutation of the SHOX gene, SRS: Silver-Russel Syndrome, and UTS: Ullrich-Turner Syndrome. An interactive version of the *Bland-Altman* plot for Deeplasia can be accessed via https://aimi-bonn.github.io/website/deeplasia/results.html and deeplasia.de.

### 3.4 Performance on longitudinal data

In clinical scenarios, determining the BA is not only important for receiving an initial diagnosis but further for monitoring the development and maturation. This requires a high test-retest reliability for the measured BA. We retrospectively estimate the test-retest reliability from regular check-ups within our cohort, employing the method proposed by Thodberg and Sävendahl (2010). In brief, this method assumes a linear progress of BA between two measurements and compares the measured BA to the interpolation between adjacent BA estimates.

The results from this analysis are summarized in Table 2 and four examples are shown in Fig. 6. Based on the GDBD we estimate the test-retest precision on patients with genetic disorders to be at most 2.74 months (95% confidence interval [2.46, 3.09], *n* = 149). Comparing our results to the ground truth rating shows that the precision of Deeplasia is on-a-par with the clinical assessment. Nevertheless, in a clinical scenario, the patient’s identity, diagnosis, and BA results from the previous examinations are known and can be used to smooth the next reported BA. If the ratings are conducted blinded and in a randomized order without additional information, the precision of the human BA reading drops significantly (Table 2) and the noise in manual BA assessment is clearly visible (Fig. 6). Thus, automatic BA prediction using Deeplasia is significantly more precise and reliable than a manual rating in a blinded scenario.

**Fig. 6.**
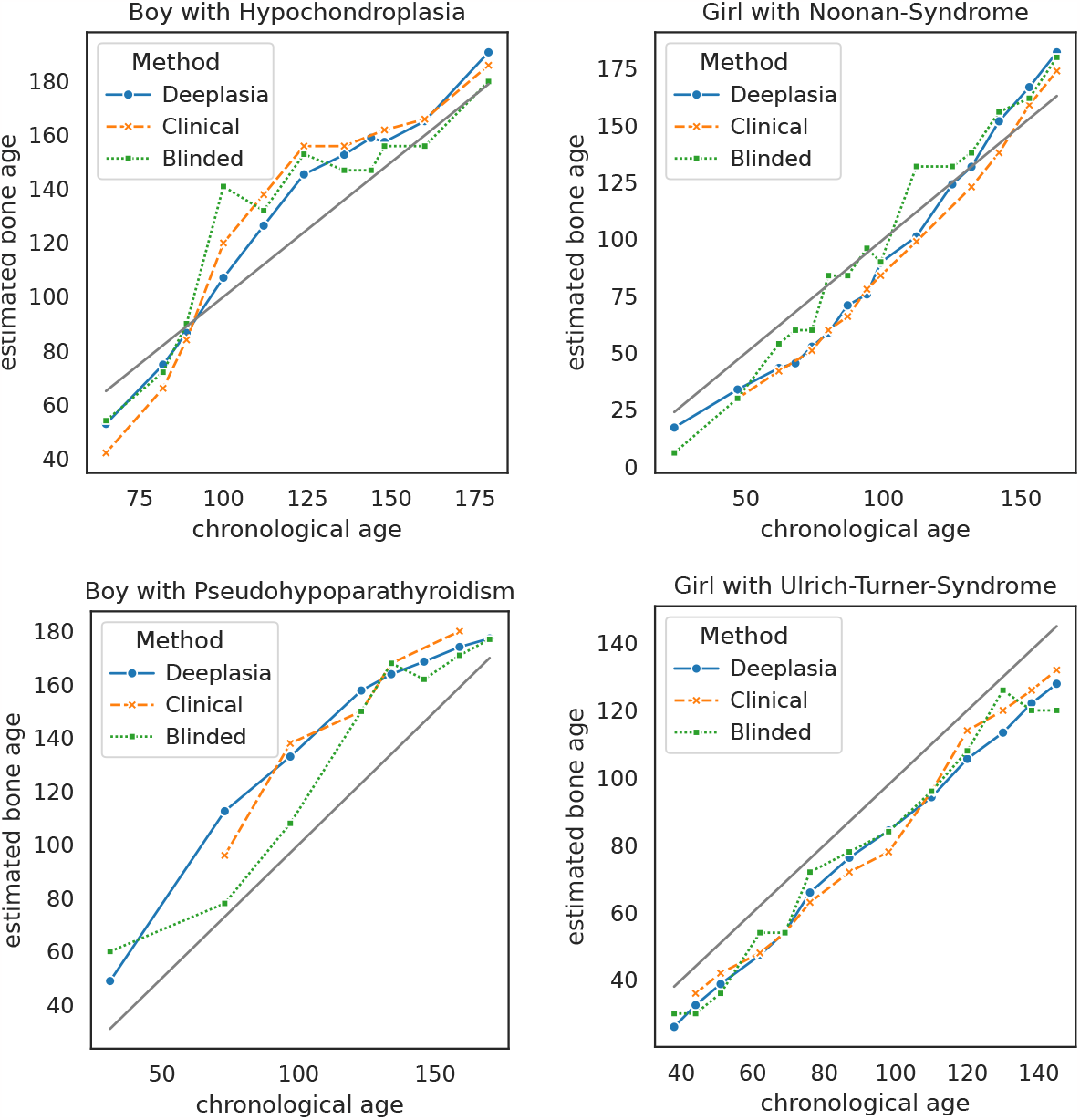
Exemplary plots of bone age maturation progress of individual patients within the GDBD estimated by Deeplasia, the clinical, and a blinded manual assessment. Bone age and chronological age are denoted in months.

## 4 Discussion

In this work, we presented Deeplasia, a deep learning approach for BA assessment specifically validated on patients with skeletal dysplasias. While designing and optimizing Deeplasia, we experimented with nine different CNN models, and to build a model ensemble, we chose three models that maximize the performance (on the validation set) and at the same time minimize the prediction patterns overlap (among the models).

With this design, Deeplasia achieves a SOTA MAD of 3.87 months on the RSNA test set, demonstrating that our prior-free learning approach is as powerful as other approaches which require additional annotations, ROI extractions, or human priors. Additionally, we tested the generalizability of Deeplasia on the external DHA dataset that contains hand X-rays of healthy patients from different ethnicities and achieved a MAD of 5.81 months. We then applied Deeplasia on the GDBD - a new dataset comprising hand X-rays with skeletal malformations - achieving a MAD of 5.96 months. These are similar to our results for the DHA set which also has two reference ratings, compared to the RSNA test set which is based on six reference ratings.

Analyzing the models’ predictive error for individual disorders, listed in Table 3, shows no significant drop in performance in comparison to the children with no diagnosed disorder. However, a tendency of increased RMSE and MAD can be observed for ACh, HyCh, and PsHPT, while a significantly decreased error can be observed for Noonan and UTS. This can likely be attributed to the accuracy of the reference grading, given that the inter-rater errors (columns 7 and 8 of Table 3) are also higher for ACh, HyCh, and, PsHPT and lower for Noonan syndrome and UTS. Of note, for each disorder, the average error of the model in comparison to a single manual rating (columns 5 and 6 of Table 3) is smaller than the average difference between the two manual raters. Hence, our model ensemble is at least as accurate as the assessed human raters for all assessed disorders and, at the same time, retains the accuracy on disorders causing severe malformations (ACh, HyCh, and, PsHPT), while those disorders increase the inter-rater disagreement.

**Table 3.**
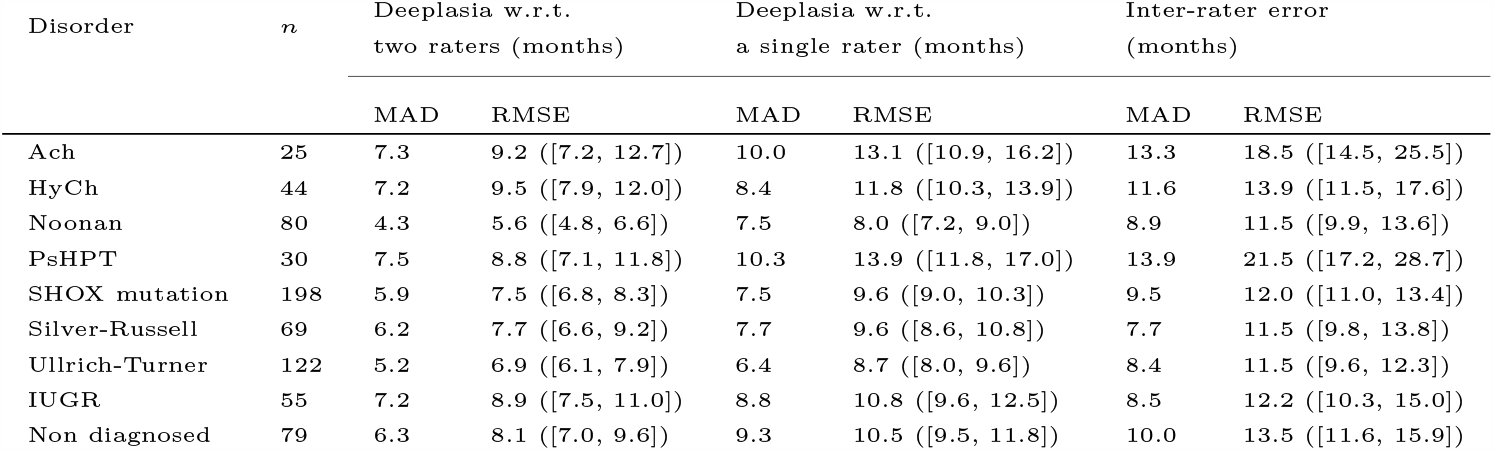
The accuracy of Deeplasia on the GDBD w.r.t. to the average bone age rating of two raters (columns 3 and 4) and a single rater (columns 5 and 6), plus the inter-rater errors (columns 7 and 8), lower MAD and RMSE errors mean higher accuracy. n refers to the number of individual X-rays per disorder. The RMSE is stated with the 95% confidence interval.

| Disorder | n | Deeplasia w.r.t.<br>two raters (months) |  | Deeplasia w.r.t.<br>a single rater (months) |  | Inter-rater error<br>(months) |  |
| --- | --- | --- | --- | --- | --- | --- | --- |
|  |  | MAD | RMSE | MAD | RMSE | MAD | RMSE |
| Ach | 25 | 7.3 | 9.2 ([7.2, 12.7]) | 10.0 | 13.1 ([10.9, 16.2]) | 13.3 | 18.5 ([14.5, 25.5]) |
| HyCh | 44 | 7.2 | 9.5 ([7.9, 12.0]) | 8.4 | 11.8 ([10.3, 13.9]) | 11.6 | 13.9 ([11.5, 17.6]) |
| Noonan | 80 | 4.3 | 5.6 ([4.8, 6.6]) | 7.5 | 8.0 ([7.2, 9.0]) | 8.9 | 11.5 ([9.9, 13.6]) |
| PsHPT | 30 | 7.5 | 8.8 ([7.1, 11.8]) | 10.3 | 13.9 ([11.8, 17.0]) | 13.9 | 21.5 ([17.2, 28.7]) |
| SHOX mutation | 198 | 5.9 | 7.5 ([6.8, 8.3]) | 7.5 | 9.6 ([9.0, 10.3]) | 9.5 | 12.0 ([11.0, 13.4]) |
| Silver-Russell | 69 | 6.2 | 7.7 ([6.6, 9.2]) | 7.7 | 9.6 ([8.6, 10.8]) | 7.7 | 11.5 ([9.8, 13.8]) |
| Ullrich-Turner | 122 | 5.2 | 6.9 ([6.1, 7.9]) | 6.4 | 8.7 ([8.0, 9.6]) | 8.4 | 11.5 ([9.6, 12.3]) |
| IUGR | 55 | 7.2 | 8.9 ([7.5, 11.0]) | 8.8 | 10.8 ([9.6, 12.5]) | 8.5 | 12.2 ([10.3, 15.0]) |
| Non diagnosed | 79 | 6.3 | 8.1 ([7.0, 9.6]) | 9.3 | 10.5 ([9.5, 11.8]) | 10.0 | 13.5 ([11.6, 15.9]) |

To have a quantitative comparison between a bone segmentation-based method with and our end-to-end approach, we applied the commonly-used BoneXpert software (Thodberg et al, 2009) on the hand X-rays contained in the GDBD. The performance of BoneXpert is shown on the right panel of Fig. 5. BoneXpert fails to assess the BA of 18 X-rays in the GDBD (11 ACh and 7 PsHPT). For the remaining 684 X-rays of the GDBD, BoneXpert achieves a MAD and an RMSE of 6.3 and 8.4 months, respectively, while (for this subset) Deeplasia achieves a MAD of 5.9 months and an RMSE of 7.6 months. The performance of BoneXpert for the individual disorders in the GDBD are listed in Table 5 in Appendix B. On the other hand, for the 18 cases rejected by BoneXpert, the MAD of Deeplasia is 9.4 months and its RMSE is 10.8 months. While there is a drop in its overall performance for these 18 cases, Deeplasia’s error is still significantly smaller than the inter-rater error (listed in Table 3). Also, as is visible in the Bland-Altman plot, Deeplasia’s predictions for these 18 cases (marked in green on the left panel of Fig. 5) show no significant deviation from the ground truth. In fact, 16 out of 18 of these cases lie within the 95% (or 1.96*σ*) confidence intervals, and the other two cases are only 2.1*σ* and 2.9*σ* from the ground truth. We remind the reader that the ground truth values are the average of two experts with in total of 60 years of experience in pediatric BA assessment. However, it would be necessary to further study the performance of Deeplasia on larger cohorts, especially to test on a larger number of ACh and PsHPT cases.

A general concern regarding medical AI is to understand its decision-making process (Amann et al, 2020). While methods relying on the segmentation of individual bones offer a higher degree of explainability compared to end-to-end learning methods, this study shows that the latter is successful in analyzing dysmorphic bones for which the former methods do not always work. However, the generalization process of the AI from normal to abnormal bones might appear difficult to comprehend. We can shed some light on the decision-making process of our end-to-end method by producing the so-called attribution maps, illustrated in Fig. 7 (see the supplementary information for details on generating these attribution maps). These maps show that the attentions of the models are mainly on the phalangeal and metacarpal joints, as well as the carpal bones, i.e. the regions relevant for BA assessment. In addition, the observable patterns in the saliency maps of the dysplastic hands remain unaltered in comparison to the hands with no diagnosed disorder. This shows that the activation patterns within the model are invariant to the dysmorphologies represented in the GDBD and the extracted features remain unaffected by the anomalies. Combined with the results of the unaltered performance, this shows the generalizability of Deeplasia to the presence of skeletal disorders in the input images.

**Fig. 7.**
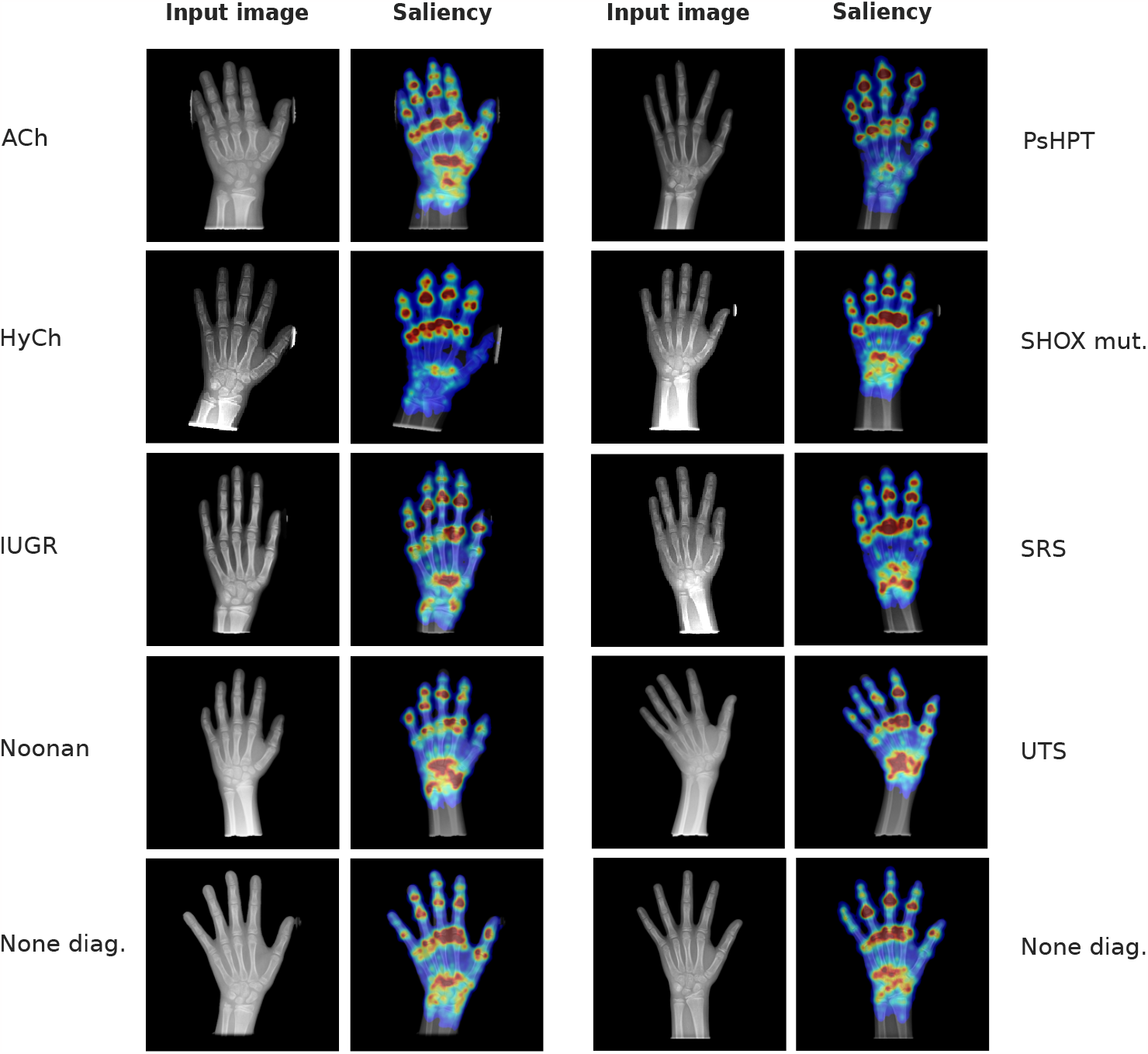
Attention heat maps from Deeplasia for bone age assessment of female patients with the indicated disorders (upper four rows) and without any diagnosed disorder from the RSNA test set (the lowest row). These maps show that the attentions of the models are mainly on the phalangeal and metacarpal joints, as well as the carpal bones, i.e. the regions relevant for BA assessment. Larger versions of these images, as well as the exact bone age measurements, are available in the supplementary information. ACh: Achondroplasia, HyCh: Hypochondroplasia, PsHPT: Pseudohypoparathyroidism, IUGR: intrauterine growth restriction, SHOX mut.: mutation of the SHOX gene, SRS: Silver-Russel Syndrome, UTS: Ullrich-Turner Syndrome, and None diag. stands for no genetic disorder diagnosed.

Finally, we applied Deeplasia to longitudinal data of patients with bone disorders. Although it only considers the X-ray images and the sex of the patients, we showed that Deeplasia is as accurate as clinical assessments (which have prior knowledge of clinical history) for monitoring the growth progress.

While there have been some studies employing DL-based techniques on medical images of patients with rare genetic diseases (e.g. Gurovich et al, 2019; Hsieh et al, 2022; Pontikos et al, 2022), this field is still understudied perhaps mainly due to the inherently small amount of available data from such diseases. The current study is limited to only seven different genetic bone diseases. Hence, our future works should expand the current dataset to a broader set of disorders and to patients with varying ethnic backgrounds^2^.

## 5 Conclusion

As patients with skeletal dysplasias are among the most important groups that require bone age assessment, it is vital to ensure the applicability and generalizability of automated approaches to these patients in dedicated studies such as this work. We demonstrated that despite being trained on non-dysplastic hand X-rays, our prior-free deep learning ensemble system, Deeplasia, generalizes to unseen bone morphologies and suits pediatric bone age assessment for skeletal dysplasias. We provide the codes we developed for our model ensemble to the community for scrutiny and reuse in their research.

## Ethics approval

The study was approved by the ethics committees of the medical faculties of the universities Magdeburg (vote 27/22) and Leipzig (vote 121/22-ek).

## Author contributions

A.K., K.S., R.G., Y.E.D.M., R.P., and K.M. collected the data. S.R., A.K. A.H., M.A.I.-A., T.-C.H., K.M., and B.J. analysed and interpreted the data. A.K., M.M.N., R.P., U.I.A., M.B., K.M., and P.M.K. provided intellectual input on clinical genetics, endocrinology, and radiology, as well as translational, ethical, and legal aspects. S.R. and B.J. designed the study, performed the statistical analysis, and drafted the initial manuscript. S.R. developed the software. B.J. conceived and supervised the study and the development of the software with input from all authors. All authors reviewed and approved the final manuscript.

## Acknowledgements

This publication has been supported by the European Reference Network on Rare Congenital Malformations and Rare Intellectual Disability (ERN-ITHACA). ERN-ITHACA is funded by the EU4Health Program of the European Union, under the Grant Agreement Nr. 101085231. The authors thank the anonymous referees for their constructive comments, Dr. Sven Koitka for the assistance with retrieving the DHA dataset and its ground truth annotations, Dr. Jörg Schaper, Dr. Alexej Knaus, Prof. Tinatin Tkemaladze, and Prof. Alain Verloes for fruitful discussions, and Dr. Hans H. Thodberg for providing access to and supporting the use of BoneXpert software as well as constructive comments on the manuscript.

## Data availability

The RSNA and the Los Angeles Digital Hand Atlas datasets are publicly available.

## Code availability

All the codes developed for training the models in this study as well as the ones used for preprocessing of the data are open-source and are publicly available at github.com/aimi-bonn/Deeplasia and github.com/aimi-bonn/hand-segmentation, respectively, or via deeplasia.de.

## Appendices

### A Predicting sex from hand X-ray and its effect on BA estimation

Biologically, bone development is highly sex-specific as girls develop and mature earlier and faster than their male peers. Consequently, the same scan read as male rather than female underestimates the BA and vice-versa (Tanner et al, 2001; Greulich and Pyle, 1959). Hence, conducting the BA assessment with the wrong sex can cause wrong results both in manual and automatic assessment (Martin et al, 2009). While such user errors are usually ignored in the model evaluation, assigning the wrong sex in a clinical setting will result in a highly inaccurate BA estimation. Yune et al (2019) demonstrated that the sex of a patient can be rather precisely predicted from a hand X-ray. Replicating their results, we integrate sex prediction into our BA estimation pipeline rendering the prediction more robust to user errors.

The sex prediction task was formulated as logistic regression. To this end, the baseline model (*EfficientNet-b0* backbone, a single FC layer of 256 neurons) was extended with an additional output neuron for the sex, while the sex was removed as an input. The model was then trained using an additional binary cross-entropy loss on the sex prediction task and the MAD was replaced by the area under the receiver operating characteristic curve (AUROC) as a validation metric.

In line with previous findings, on the RSNA test set our sex prediction model achieves an accuracy of 93.0%, 89.3%, and 81.8% for the RSNA test set, the DHA, and the GDBD, respectively. Using the sex predicted by the model as input to our bone age models, the accuracy in each test set drops considerably (Table 4). Hence, completely omitting sex annotated by the user would result in a dramatic loss of accuracy. Therefore, we propose to use the sex prediction as mainly a verification step to mark contradictions between user input and model prediction followed by the user double-checking to potentially correct erroneous inputs.

**Table 4.**
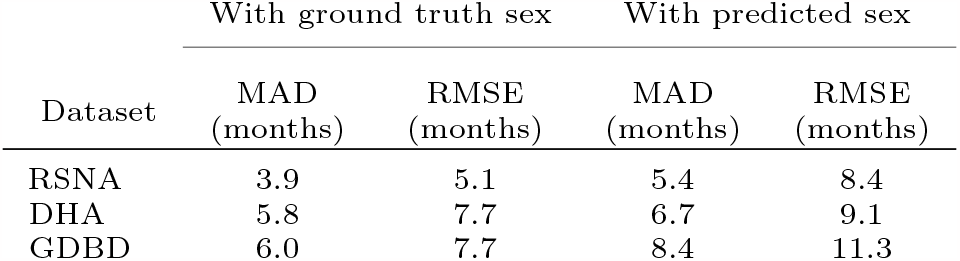
Performance of the model ensemble on different datasets using either the real biological sex (ground truth) or the sex predicted by the dedicated model.

### B Comparison with BoneXpert

Table 5 lists the performance of BoneXpert in the BA assessment of different disorders in the GDBD. The performance of Deeplasia is also listed again for quick comparison. BoneXpert rejected 11 out of 25 (44%) of Ach cases and 7 out of 30 (23%) of PsHPT cases. The BoneXpert rejection rate for Ach is in agreement with the expected ≈ 50% (personal communication with H. H. Thodberg, March 2023). BoneXpert performs better for cases with HyCh, Silver-Russel syndrome, and IUGR. The performance of both software is similar for Noonan and (the non-rejected) PsHPT cases. On the other hand, Deeplasia performs better in cases with SHOX mutation and Ulrich-Turner syndrome, and significantly better for cases with Achondroplasia.

**Table 5.**
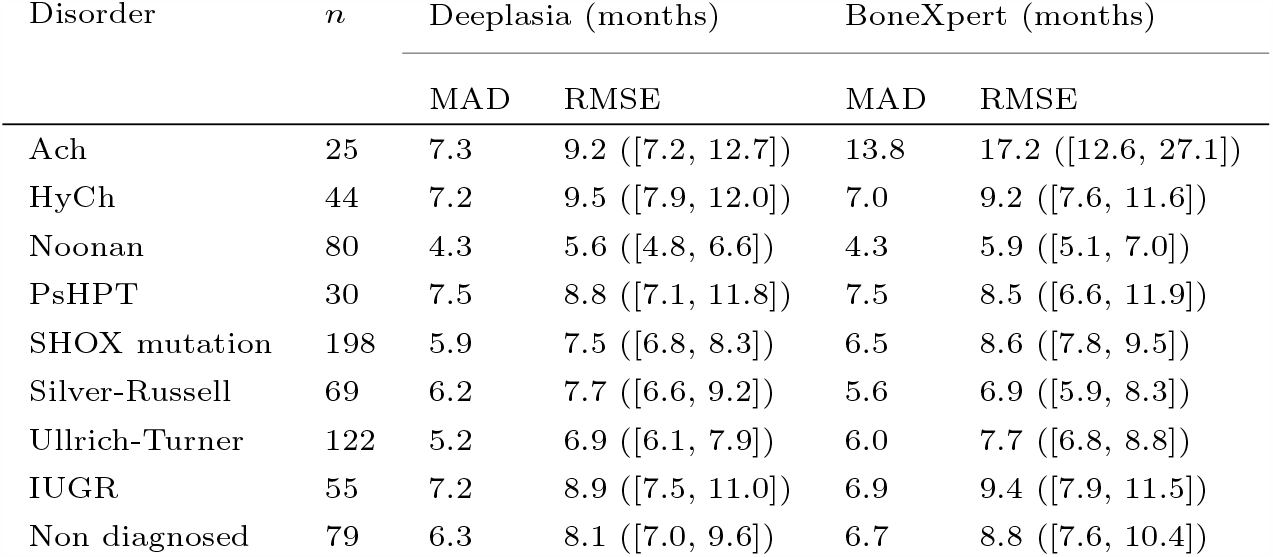
Comparing the performance of Deeplasia and BoneXpert in the BA assessment of different disorders in the GDBD. Lower MAD and RMSE errors mean higher accuracy. The RMSE is stated with the 95% confidence interval. n refers to the number of individual X-rays per disorder. BoneXpert rejected 11 out of 25 (44%) of Ach cases and 7 out of 30 (23%) of PsHPT cases.

## Footnotes

1 We further discuss the effect of patient’s sex on BA assessment in Appendix A.

2 E.g. via support from FAIR (Wilkinson et al, 2016) sources such as the [GestaltMatcherDatabase](https://db.gestaltmatcher.org/) (Lesmann et al, 2023).

## References

Aicardi G, Vignolo M, Milani S, et al (2000) Assessment of skeletal maturity of the hand-wrist and knee: A comparison among methods. Am J Hum Biol 12(5):610–615

Amann J, Blasimme A, et al (2020) Explainability for artificial intelligence in healthcare: a multidisciplinary perspective. BMC Medical Informatics and Decision Making 20(1). 10.1186/s12911-020-01332-6, URL 10.1186/s12911-020-01332-6

Bunch PM, Altes TA, McIlhenny J, et al (2017) Skeletal development of the hand and wrist: digital bone age companion-a suitable alternative to the greulich and pyle atlas for bone age assessment? Skeletal Radiol 46(6):785–793

Cicero M, Bilbily A (2017) Machine learning and the future of radiology: How we won the 2017 rsna ml challenge. Accessed from https://www16bitai/blog/ml-and-future-of-radiology

Creo AL, Schwenk WF2nd (2017) Bone age: A handy tool for pediatric providers. Pediatrics 140(6)

De Sanctis V, Di Maio S, Soliman AT, et al (2014) Hand x-ray in pediatric endocrinology: Skeletal age assessment and beyond. Indian J Endocrinol Metab 18(Suppl 1):S63–71

Eng DK, Khandwala NB, Long J, et al (2021) Artificial intelligence algorithm improves radiologist performance in skeletal age assessment: A prospective multicenter randomized controlled trial. Radiology 301(3):692–699

Escobar M, González C, Torres F, et al (2019) Hand pose estimation for pediatric bone age assessment. In: Medical Image Computing and Computer Assisted Intervention – MICCAI 2019. Springer International Publishing, pp 531–539

Gertych A, Zhang A, Sayre J, et al (2007) Bone age assessment of children using a digital hand atlas. Computerized medical imaging and graphics 31(4-5):322–331

Greulich WW, Pyle SI (1959) Radiographic Atlas of Skeletal Development of the Hand and Wrist. Stanford University Press

Gurovich Y, Hanani Y, Bar O, et al (2019) Identifying facial phenotypes of genetic disorders using deep learning. Nature Medicine 25(1):60–64. 10.1038/s41591-018-0279-0, URL 10.1038/s41591-018-0279-0

Halabi SS, Prevedello LM, Kalpathy-Cramer J, et al (2019) The RSNA pediatric bone age machine learning challenge. Radiology 290(2):498–503

Hsieh TC, Bar-Haim A, Moosa S, et al (2022) GestaltMatcher facilitates rare disease matching using facial phenotype descriptors. Nature Genetics 54(3):349–357. 10.1038/s41588-021-01010-x, URL https://doi.org/10.1038/s41588-021-01010-x

Kim JR, Shim WH, Yoon HM, et al (2017) Computerized bone age estimation using deep learning based program: Evaluation of the accuracy and efficiency. AJR Am J Roentgenol 209(6):1374–1380

Koitka S, Kim MS, Qu M, et al (2020) Mimicking the radiologists’ workflow: Estimating pediatric hand bone age with stacked deep neural networks. Medical Image Analysis 64:101,743. 10.1016/j.media.2020.101743, URL https://doi.org/10.1016/j.media.2020.101743

Larson DB, Chen MC, Lungren MP, et al (2018) Performance of a Deep-Learning neural network model in assessing skeletal maturity on pediatric hand radiographs. Radiology 287(1):313–322

Lesmann H, Lyon GJ, Caro P, et al (2023) Gestaltmatcher database - a fair database for medical imaging data of rare disorders. medRxiv 10.1101/2023.06.06.23290887, URL https://www.medrxiv.org/content/early/2023/06/10/2023.06.06.23290887, https://arxiv.org/abs/ https://www.medrxiv.org/content/early/2023/06/10/2023.06.06.23290887.full.pdf

Martin DD, Deusch D, Schweizer R, et al (2009) Clinical application of automated greulich-pyle bone age determination in children with short stature. Pediatric radiology 39(6):598–607

Martin DD, Wit JM, Hochberg Z, et al (2011a) The use of bone age in clinical practice–part 1. Hormone research in paediatrics 76(1):1–9

Martin DD, Wit JM, Hochberg Z, et al (2011b) The use of bone age in clinical practice–part 2. Hormone research in paediatrics 76(1):10–16

Martin DD, Calder AD, Ranke MB, et al (2022) Accuracy and self-validation of automated bone age determination. Sci Rep 12(1):6388

Offiah AC (2021) Current and emerging artificial intelligence applications for pediatric musculoskeletal radiology. Pediatric Radiology 52(11):2149–2158. 10.1007/s00247-021-05130-8, URL https://doi.org/10.1007%2Fs00247-021-05130-8

Offiah AC, Hall CM (2020) The radiologic diagnosis of skeletal dysplasias: past, present and future. Pediatr Radiol 50(12):1650–1657

Pontikos N, Woof W, Veturi A, et al (2022) Eye2gene: prediction of causal inherited retinal disease gene from multimodal imaging using deep-learning 10.21203/rs.3.rs-2110140/v1, URL https://doi.org/10.21203/rs.3.rs-2110140/v1

Rajpurkar P, Chen E, Banerjee O, et al (2022) AI in health and medicine. Nature Medicine 28(1):31–38. 10.1038/s41591-021-01614-0, URL 10.1038%2Fs41591-021-01614-0

Rassmann S, Hustinx A, Krawitz PM, et al (2023) Hand mask for the rsna bone age dataset. 10.5281/zenodo.7611677, URL https://doi.org/10.5281/zenodo.7611677

Sabir AH, Cole T (2019) The evolving therapeutic landscape of genetic skeletal disorders. Orphanet Journal of Rare Diseases 14(1). 10.186/s13023-019-1222-2, URL https://doi.org/10.1186/s13023-019-1222-2

Satoh M, Hasegawa Y (2022) Factors affecting prepubertal and pubertal bone age progression. Front Endocrinol 13:967,711

Spranger J, Superti-Furga A, Unger S (2018) Bone Dysplasias: An Atlas of Genetic Disorders of Skeletal Development. Oxford University Press, URL https://books.google.de/books?id=jLhwDwAAQBAJ

Tan M, Le Q (2019) Efficientnet: Rethinking model scaling for convolutional neural networks. In: International conference on machine learning, PMLR, pp 6105–6114

Tanner JM (1962) Growth at adolescence; with a general consideration of the effects of hereditary and environmental factors upon growth and maturation from birth to maturity. Blackwell Scientific Publications, Oxford

Tanner JM, Whitehouse RH, Cameron N, et al (1975) Assessment of skeletal maturity and prediction of adult height (TW2 method)

Tanner JM, Healy MJR, Goldstein H, et al (2001) Assessment of skeletal maturity and prediction of adult height (TW3 method), 3rd edn. W.B. Saunders, London

Thodberg HH, Sävendahl L (2010) Validation and reference values of automated bone age determination for four ethnicities. Academic radiology 17(11):1425–1432

Thodberg HH, Kreiborg S, Juul A, et al (2009) The BoneXpert method for automated determination of skeletal maturity. IEEE Trans Med Imaging 28(1):52–66

Torres F, González C, Escobar MC, et al (2020) An empirical study on global bone age assessment. In: 15th International Symposium on Medical Information Processing and Analysis, SPIE, pp 98–105

Unger S, Ferreira CR, Mortier GR, et al (2023) Nosology of genetic skeletal disorders: 2023 revision. American Journal of Medical Genetics Part A 191(5):1164–1209. 10.1002/ajmg.a.63132, URL https://onlinelibrary.wiley.com/doi/abs/10.1002/ajmg.a.63132, https://arxiv.org/abs/https://onlinelibrary.wiley.com/doi/pdf/10.1002/ajmg.a.63132

Wang D, Zhang K, Ding J, et al (2020a) Improve bone age assessment by learning from anatomical local regions. In: International Conference on Medical Image Computing and Computer-Assisted Intervention, Springer, pp 631–640

Wang F, Gu X, Chen S, et al (2020b) Artificial intelligence system can achieve comparable results to experts for bone age assessment of chinese children with abnormal growth and development. PeerJ 8:e8854

Wilkinson MD, Dumontier M, Aalbersberg IJ, et al (2016) The FAIR guiding principles for scientific data management and stewardship. Scientific Data 3(1). 10.1038/sdata.2016.18, URL https://doi.org/10.1038/sdata.2016.18

Yune S, Lee H, Kim M, et al (2019) Beyond human perception: sexual dimorphism in hand and wrist radiographs is discernible by a deep learning model. Journal of Digital Imaging 32(4):665–671

Zech JR, Badgeley MA, Liu M, et al (2018) Variable generalization performance of a deep learning model to detect pneumonia in chest radiographs: a cross-sectional study. PLoS medicine 15(11):e1002.683

Zhang A, Sayre JW, Vachon L, et al (2009) Racial differences in growth patterns of children assessed on the basis of bone age. Radiology 250(1):228–235

