## Supplementary Information for "Deeplasia: prior-free deep learning for pediatric bone age assessment robust to skeletal dysplasias"

### Supplementary Material 1: Hand segmentation

To extract the hand and mask the background, masks representing the hand were annotated on 528 images randomly drawn from the RSNA training set. The mask annotation used a semi-automatic procedure based on applying intensity thresholds and edge detection. The segmentation was manually controlled and, if needed, corrected. Leveraging this dataset, we trained *TensorMask* (Chen et al, 2019) and *Efficient-UNet* models (Baheti et al, 2020) for automated mask prediction using 460 masks and leaving the remaining masks for validation. To avoid fitting the BA model to the masks predicted by only one of the masking models and potentially wrongly predicted masks decreasing the effective size of the training set, in each training epoch the masks were randomly selected between either model. To allow for fast mask prediction without hardware acceleration, a light-weight *FastSurferCNN* (Henschel et al, 2020) model was trained on the masks predicted by the *TensorMask* model of the complete RSNA BA training set excluding images with manually edited masks. To reduce the model size, the number of filters was reduced to 32. The models were trained using the ADAM optimizer (Kingma and Ba, 2014) with a base learning rate (LR) of $10^{-2}$ scheduled with the *CosineAnnealingLR* ($T_0 = 10$, minimum LR of $5 \cdot 10^{-5}$) for 50 epochs and a batch size of 32. Weight decay was set to $10^{-6}$. To reduce biases towards detecting all high-intensity pixels as false positive, the weight of the logistic loss term of the composite loss (Henschel et al, 2020) was adapted to also include all pixels outside of the annotated mask with an intensity higher than the 80<sup>th</sup> percentile of the annotated mask. All images were standardized to a mean intensity of zero and a standard deviation of one. We chose this standardization over the commonly used min-max normalization as the latter would be highly susceptible to few

high-intensity pixels e.g. due to remaining scanning or imaging artifacts. Further, we simulated scanning artifacts by drawing artificial boxes and gradient stripes. With this configuration, we achieved a Dice similarity score of 0.993 with respect to the unseen manually edited set of masks. To test the performance of the full BA prediction pipeline, the model evaluation was carried out using *FastSurferCNN* masking.

### Supplementary Material 2: Details of bone age model training

All BA models were trained using a mean squared error (MSE) loss and the ADAM optimizer (Kingma and Ba, 2014). The initial LR was set to $10^{-3}$ and decayed using *ReduceLROnPlateau* (factor of 0.2, patience of 10 epochs) to a minimum LR of $10^{-4}$ tracking the MAD in the validation set. The weights of the final models from each training process were chosen based on the best validation MAD (“checkpointing”). For regularization, dropout ($p = 0.2$) was added to the FC layers and a weight decay of $5 * 10^{-6}$ was applied. We resized the images to a minimum resolution of $512 \times 512$ to assert the resoluting potentially relevant fine-grained structures in the bones such as growth gaps. Due to the miss-match of the training resolution with the pre-training of the smaller *EfficientNet-b0* version, the *-b0* models were trained from scratch using the Kaiming initialization (He et al, 2015). However, the larger *-b4* versions are pre-trained at a similar resolution, so the *-b4* models used *ImageNet* (Deng et al, 2009) pre-training. The *-b0* versions were trained for 300 epochs, whereas the pre-training of the *-b4* models allowed for faster convergence, so the training was reduced to 100 epochs. The mini-batch size was set to 32 for all models.

As *default* data augmentations we used the approach described by Cicero and Bilbily (2017) (relative scaling and translation of $\pm 0 - 20\%$, rotation of $\pm 0 - 20^\circ$, shear of $\pm 0 - 1^\circ$ horizontal flipping with $p = 0.5$). We extended this to our *strong* set of augmentations by increasing the maximum scaling and translation to $\pm 30\%$, rotation to $\pm 30^\circ$, and shear to $\pm 10^\circ$. Additionally, non-linear intensity manipulations with either ($p = 0.33$) a Gamma-correction (gamma chosen from $[0.7, 1.3]$) or ($p = 0.67$) a contrast limited adaptive histogram equalization (CLAHE, Pizer et al, 1987, clip limit: 3), and image sharpening ($p = 0.2$, alpha chosen from $[0.5, 0.75]$ and lightness chosen from $[0.5, 1]$) were applied. To compensate for strong regularization inducing a bias towards predicting more extreme BAs on non-augmented samples, the inferred predictions were corrected via a linear regression model fitted on the predictions of the non-augmented training set.

Test time augmentation (TTA) was performed by rotating the input image by $-10, -5, 0, 5, 10^\circ$ and each with and without applying additional horizontal mirroring. Both, model ensembling and TTA, use an unweighted average of all predictions for any given image.

The models included in our final ensemble were chosen based on the best validation MAD score in each training condition. Figure 1 of this supplementary information shows a sketch of the model architecture. The models were implemented in *PyTorch* (v1.10, Paszke et al, 2019) using the *lightning* framework (v1.6, Falcon et al, 2019). We used the *Detectron2* (v0.4, Wu et al, 2019) implementation of *Tensormask* Chen et al (2019). Data augmentation was conducted using the *Albumentations* library (v1.1, Buslaev et al, 2020). Image pre- and postprocessing was conducted in *OpenCv* (v4.5, Bradski, 2000).

### **Predicting sex from hand X-ray and its effect on BA estimation**

Biologically, bone development is highly sex-specific as girls develop and mature earlier and faster than their male peers. Consequently, the same scan read as male rather than female underestimates the BA and vice-versa (Tanner et al, 2001; Greulich and Pyle, 1959). Hence, conducting the BA assessment with the wrong sex can cause wrong results both in manual and automatic assessment (Martin et al, 2009). While such user errors are usually ignored in the model evaluation, assigning the wrong sex in a clinical setting will result in a highly inaccurate BA estimation. Yune et al (2019) demonstrated that the sex of a patient can be rather precisely predicted from a hand X-ray. Replicating their results, we integrate sex prediction into our BA estimation pipeline rendering the prediction more robust to user errors. The sex prediction task was formulated as logistic regression. To this end, the baseline model (EfficientNet-b0 backbone, a single FC layer of 256 neurons) was extended with an additional output neuron for the sex, while the sex was removed as an input. The model was then trained using an additional binary cross-entropy loss on the sex prediction task and the MAD was replaced by the area under the receiver operating characteristic curve (AUROC) as a validation metric. In line with previous findings, on the RSNA test set our sex prediction model achieves an accuracy of 93.0%, 89.3%, and 81.8% for the RSNA test set, the Digital Hand Atlas, and the German Dysplastic Bone Dataset, respectively. Using the sex predicted by the model as input to our bone age models, the accuracy in each test set drops considerably (Table 1). Hence, completely omitting sex annotated by the user would result in a dramatic loss of accuracy.

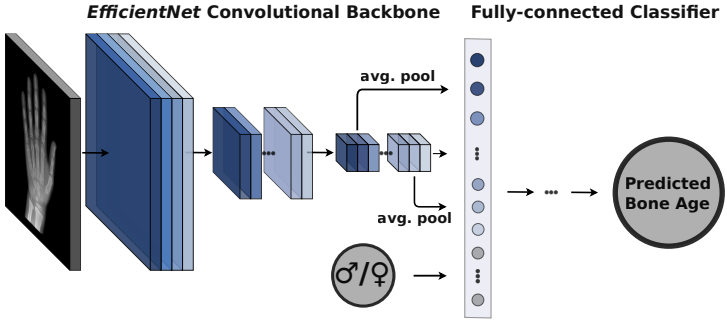

**Fig. 1** Model architecture for bone age prediction. The gray-scale input image is passed through an *EfficientNet* backbone model. The obtained features are combined with an inflated representation of the sex and passed into a fully-connected network to obtain the bone age.

Therefore, we propose to use the sex prediction as mainly a verification step to mark contradictions between user input and model prediction followed by the user double-checking to potentially correct erroneous inputs.

| Dataset | With ground truth sex |  | With predicted sex |  |
| --- | --- | --- | --- | --- |
|  | MAD<br>(months) | RMSE<br>(months) | MAD<br>(months) | RMSE<br>(months) |
| RSNA | 3.9 | 5.1 | 5.4 | 8.4 |
| DHA | 5.8 | 7.7 | 6.7 | 9.1 |
| GDBD | 6.0 | 7.7 | 8.4 | 11.3 |

**Table 1** Performance of the model ensemble on different datasets using either the real biological sex (ground truth) or the sex predicted by the dedicated model.

### Supplementary Material 3: Model experimentations

DL model ensembles often show higher performances compared to single models (see e.g. Pan et al, 2019; Hustinx et al, 2023; Pontikos et al, 2022), however, usually multiple experimentations are required to reach a suitable set of models. We took the following steps to investigate the optimum model configurations:

In the first experiment, the effect of applying a stronger data augmentation than previously proposed by Cicero and Bilbily (2017) was studied. To this end, we compared the performance of the smallest model configurations (*EfficientNet-b0* with $512 \times 512$ input resolution) trained with *default* and *strong* augmentations. These include additional non-linear intensity transformations and edge sharpening (see the previous section of this supplementary information).

Assessing the performance of these models on the internal validation set of the RSNA dataset shows that the *strong* augmentations improve the prediction accuracy across all assessed model configurations (see Table 2 of this supplementary information for details). Therefore, we assumed that the *strong* augmentations would improve generalization to unseen data and used these augmentations for all the subsequent experiments as our *baseline* training condition.

Next, we studied the effect of scaling the model size by replacing the *EfficientNet-b0* backbone ($5.3 * 10^6$ parameters) with the larger *-b4* version ($19.3 * 10^6$ parameters) in the *large CNN* condition. Finally, we explored the effect of increasing the input resolution from $512 \times 512$ to $1024 \times 1024$ as the

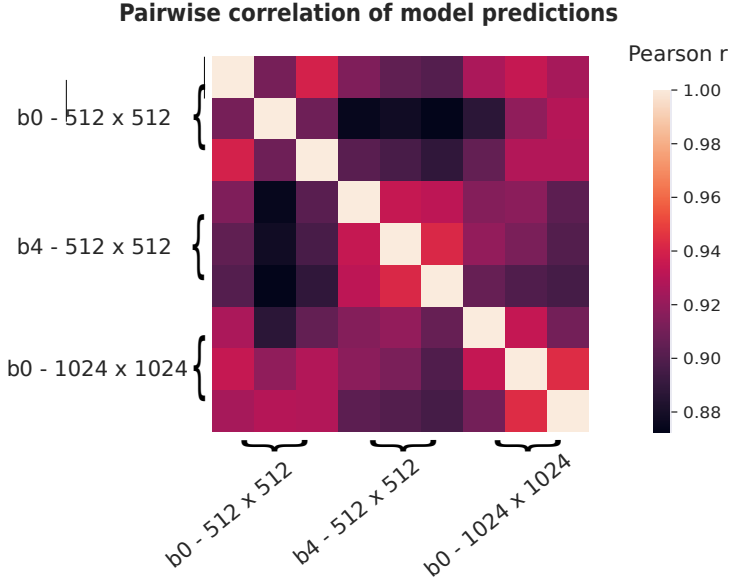

**Fig. 2** Pairwise correlations of the predicted bone ages (BAs) on the RSNA validation dataset of nine models with different *EfficientNet* backbone models (*EfficientNet-b0* and *-b4*) and at different image resolutions ($512 \times 512$ and $1024 \times 1024$). For each combination of backbone and resolution models with various sets of FC layers ([256], [512, 512], [1024, 1024, 512, 512], top to bottom / left to right) were trained and validated. The correlation of the predicted BAs is stated as Pearson’s correlation coefficient.

*high-resolution* condition. Both of the latter modifications show additional improvements compared to the baseline configuration in the RSNA validation set (Table 3 of this supplementary information).

As a final experiment, we conducted test time augmentation (TTA) similar to the method proposed by Cicero and Bilbily (2017) for the three chosen models. This improved the ensemble validation accuracy MAD from 6.12 to 6.08 months, see Table 4 of this supplementary information. Given that the TTA yielded only a marginal improvement but has high computational costs, we decided not to include it in our BA inference approach.

The Tables 2, 3, and 4 of this supplementary information list the detailed results of our experimentation for building the model ensemble.

| *EfficientNet*<br>version | Input resolution | FC layers | Val. MAD (months) |  |
| --- | --- | --- | --- | --- |
|  |  |  | *default* augm. | *strong* augm. |
| *b0* | $512 \times 512$ | [256] | 6.6 | 6.4 |
| *b0* | $512 \times 512$ | [512, 512] | 6.8 | 6.7 |
| *b0* | $512 \times 512$ | [1024, 1024, 512, 512] | 6.5 | 6.4 |

**Table 2** Accuracy of single models trained with the *default* and *strong* set of augmentations at different configurations of fully-connected (FC) layers.

| Condition name | *EfficientNet*<br>version | Input resolution | FC layers | Val. MAD (months) |
| --- | --- | --- | --- | --- |
| *baseline* | *b0* | $512 \times 512$ | [256] | 6.4* |
|  |  |  | [512, 512] | 6.7 |
|  |  |  | [1024, 1024, 512, 512] | 6.4 |
| *large CNN* | *b4* | $512 \times 512$ | [256] | 6.3 |
|  |  |  | [512, 512] | 6.4* |
|  |  |  | [1024, 1024, 512, 512] | 6.4 |
| *high-resolution* | *b0* | $1024 \times 1024$ | [256] | 6.3* |
|  |  |  | [512, 512] | 6.4 |
|  |  |  | [1024, 1024, 512, 512] | 6.4 |

**Table 3** Comparison of the validation MAD of different training conditions and model configurations. The final model ensemble was selected based on the best score (marked by \* in this table) in each training condition.

| Condition name | *EfficientNet*<br>version | Input resolution | FC layers | Val. MAD (months) |  |
| --- | --- | --- | --- | --- | --- |
|  |  |  |  | no TTA | TTA |
| *baseline* | *b0* | $512 \times 512$ | [256] | 6.4 | 6.4 |
| *high-resolution* | *b0* | $1024 \times 1024$ | [256] | 6.3 | 6.2 |
| *large CNN* | *b4* | $512 \times 512$ | [512, 512] | 6.2 | 6.1 |
| ensemble |  |  |  | 6.1 | 6.1 |

**Table 4** Comparison of the best performing single models in each condition on the RSNA BA validation set with and without applying test time augmentation (TTA). Additionally, an ensemble composed of all models is tested.

### Supplementary Material 4: Attention maps

The attribution maps $M$ were generated by calculating the absolute value of the gradient of the predicted BA $\hat{Y}$ w.r.t. to the input image $I$ given sex $S$:

$$M(I) = \left| \frac{\partial \hat{Y}}{\partial I'} \right|_{I'=I} = \left| \frac{\partial f^{(w)}(I, S)}{\partial I'} \right|_{I'=I} \quad (1)$$

The resulting image was subsequently smoothed using a Gaussian kernel with a size 5% of the input image resolution. Subsequently, the maps were normalized by subtracting the minimum intensity, dividing by value of the resulting 99<sup>th</sup> percentile, and clipping all resulting values to a maximum of 1. For better visualization of the results in the scenario of masked input images, values less than 0.075 were excluded before applying the color map. Finally, the color maps were blended on the original input images.

Figures 3-7 of this supplementary information show the large versions of the selected hand X-rays presented in Figure 8 of the main manuscript with the estimated bone ages and the attention maps from Deeplasia.

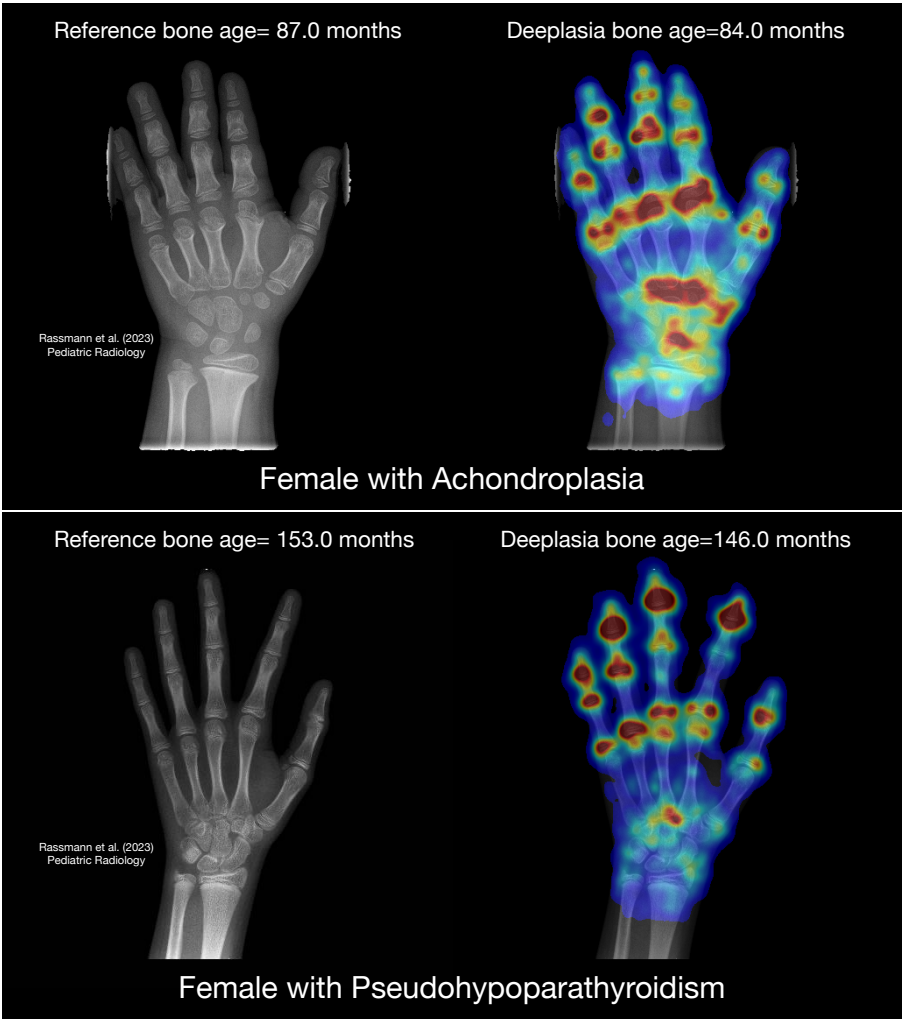

**Fig. 3** Attention heat maps from Deeplasia.

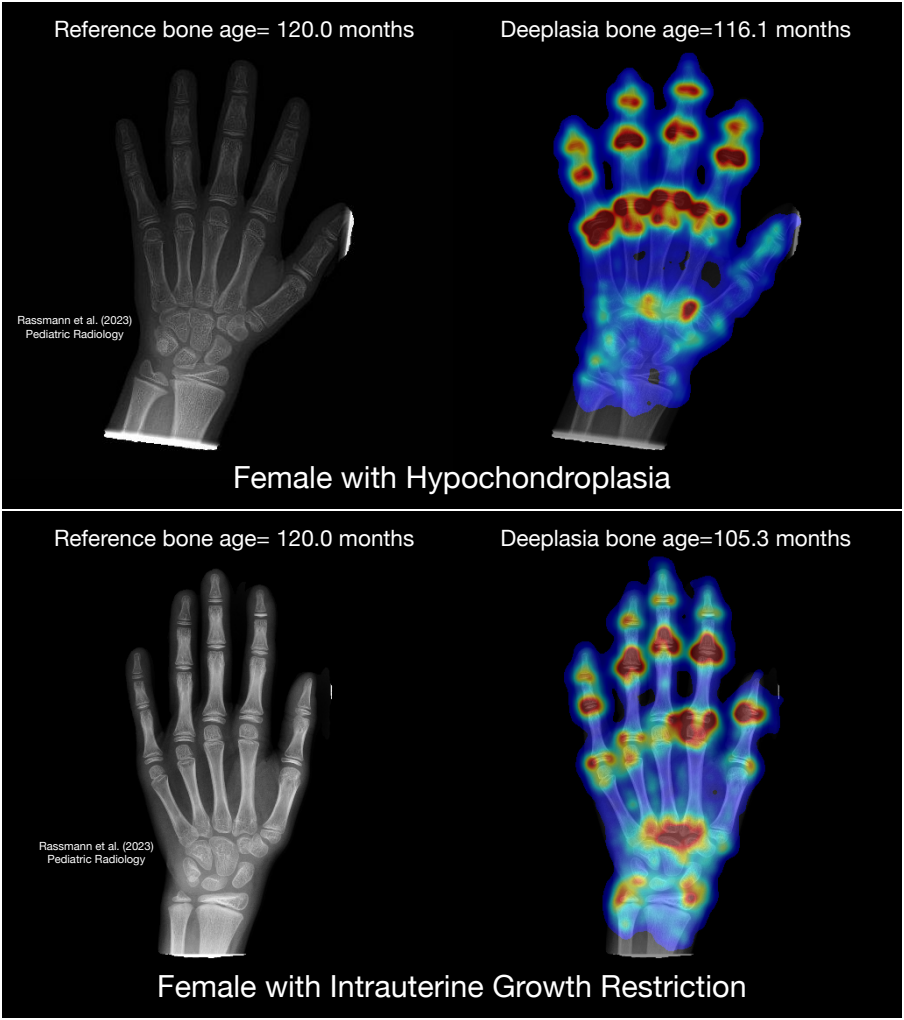

**Fig. 4** Attention heat maps from Deeplasia.

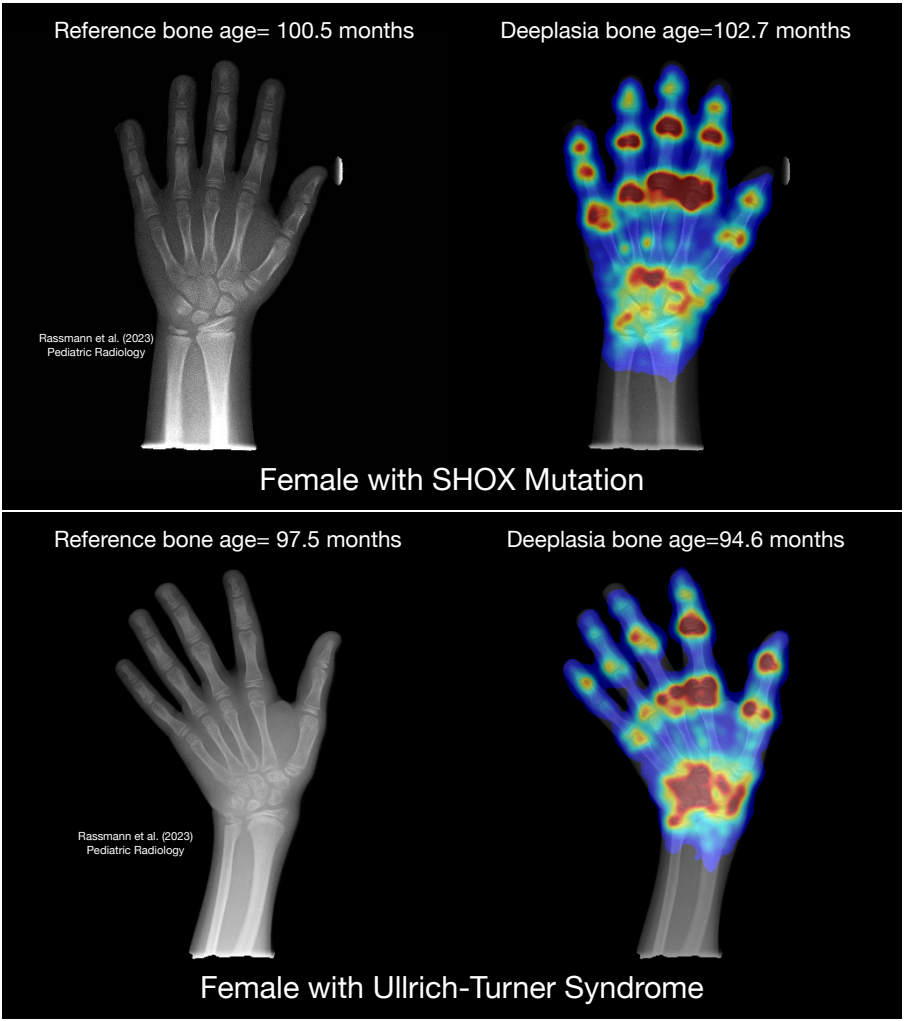

**Fig. 5** Attention heat maps from Deeplasia.

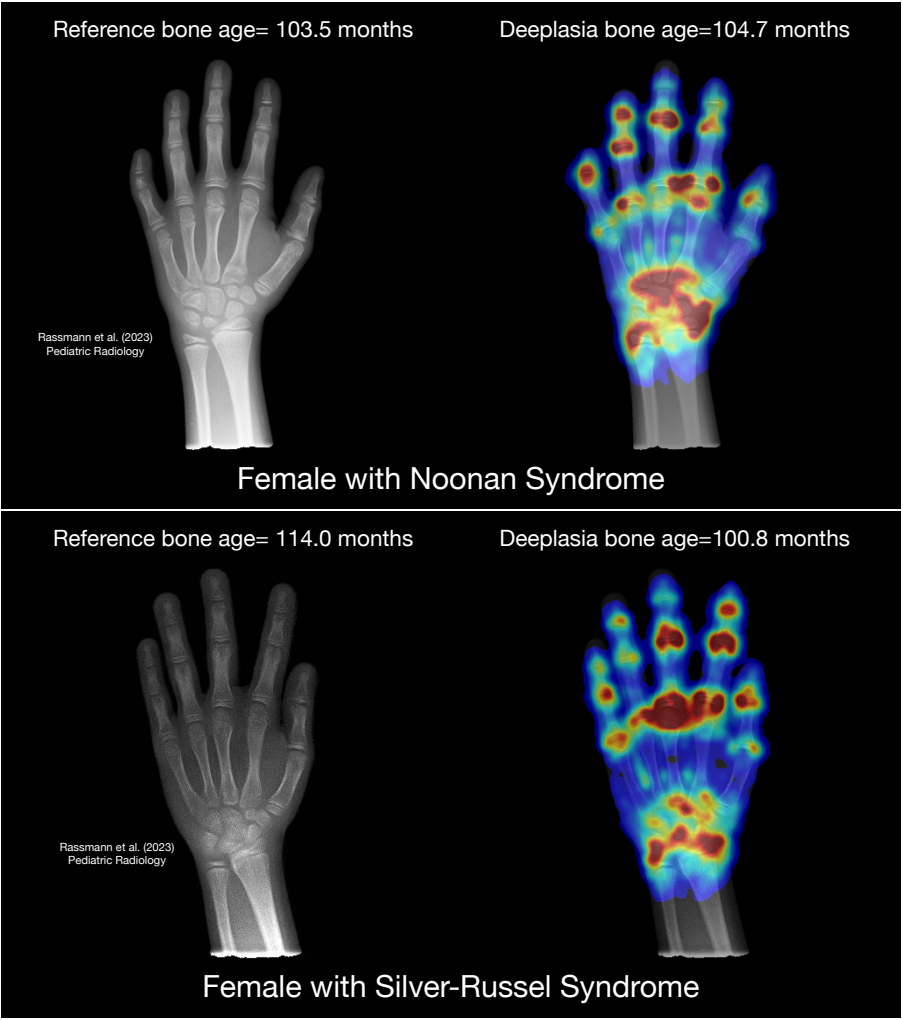

**Fig. 6** Attention heat maps from Deeplasia.

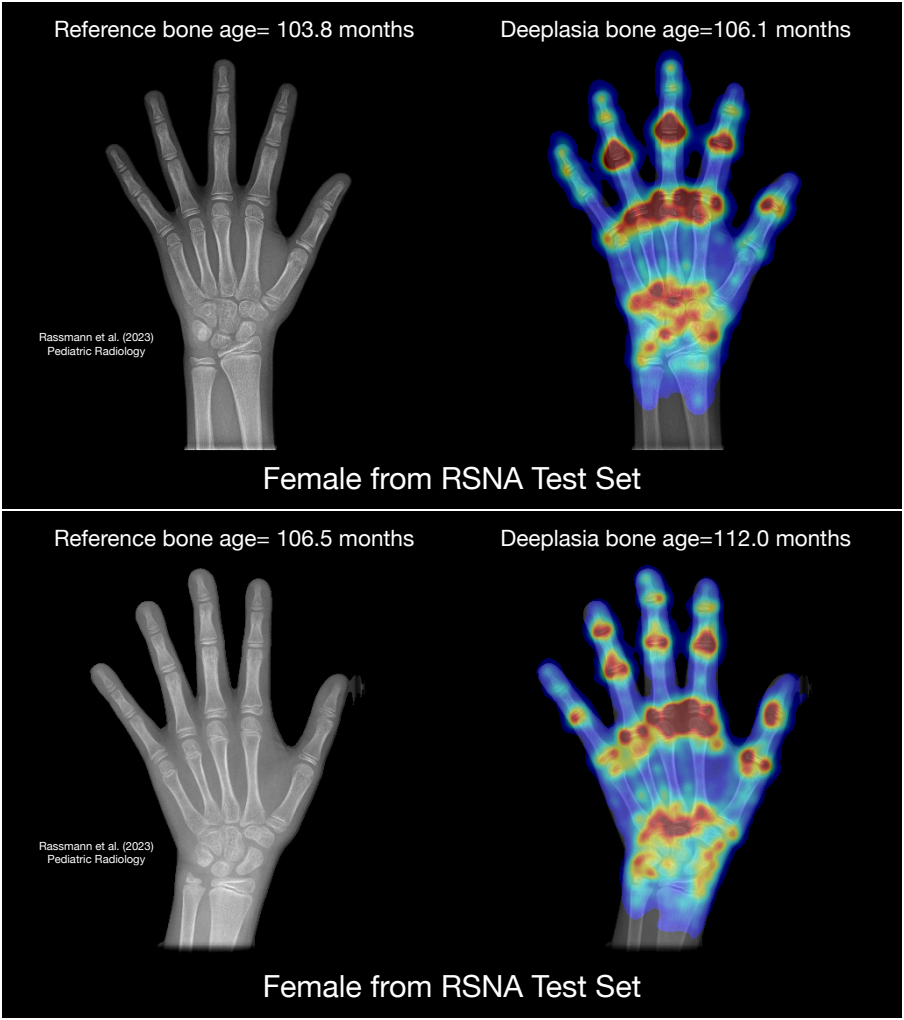

**Fig. 7** Attention heat maps from Deeplasia.

### Supplementary Material 5: Comparison with BoneXpert

Table 5 lists the performance of BoneXpert in the BA assessment of different disorders in the German Dysplastic Bone Dataset. The performance of Deeplasia is also listed again for quick comparison. BoneXpert rejected 11 out of 25 (44%) of achondroplasia cases and 7 out of 30 (23%) of pseudohypoparathyroidism cases. The BoneXpert rejection rate for achondroplasia is in agreement with the expected $\approx 50\%$ (personal communication with H. H. Thodberg, March 2023). BoneXpert performs better for cases with hypochondroplasia, Silver-Russell syndrome, and IUGR. The performance of both software is similar for Noonan and (the non-rejected) pseudohypoparathyroidism cases. On the other hand, Deeplasia performs better in cases with SHOX mutation and Ulrich-Turner syndrome, and significantly better for cases with achondroplasia.

| Disorder | *n* | Deeplasia (months) |  | BoneXpert (months) |  |
| --- | --- | --- | --- | --- | --- |
|  |  | MAD | RMSE | MAD | RMSE |
| Ach | 25 | 7.3 | 9.2 ([7.2, 12.7]) | 13.8 | 17.2 ([12.6, 27.1]) |
| HyCh | 44 | 7.2 | 9.5 ([7.9, 12.0]) | 7.0 | 9.2 ([7.6, 11.6]) |
| Noonan | 80 | 4.3 | 5.6 ([4.8, 6.6]) | 4.3 | 5.9 ([5.1, 7.0]) |
| PsHPT | 30 | 7.5 | 8.8 ([7.1, 11.8]) | 7.5 | 8.5 ([6.6, 11.9]) |
| SHOX mutation | 198 | 5.9 | 7.5 ([6.8, 8.3]) | 6.5 | 8.6 ([7.8, 9.5]) |
| Silver-Russell | 69 | 6.2 | 7.7 ([6.6, 9.2]) | 5.6 | 6.9 ([5.9, 8.3]) |
| Ullrich-Turner | 122 | 5.2 | 6.9 ([6.1, 7.9]) | 6.0 | 7.7 ([6.8, 8.8]) |
| IUGR | 55 | 7.2 | 8.9 ([7.5, 11.0]) | 6.9 | 9.4 ([7.9, 11.5]) |
| Non diagnosed | 79 | 6.3 | 8.1 ([7.0, 9.6]) | 6.7 | 8.8 ([7.6, 10.4]) |

**Table 5** Comparing the performance of Deeplasia and BoneXpert in the BA assessment of different disorders in the GDBD. Lower MAD and RMSE errors mean higher accuracy. The RMSE is stated with the 95% confidence interval. *n* refers to the number of individual radiographs per disorder. BoneXpert rejected 11 out of 25 (44%) of achondroplasia cases and 7 out of 30 (23%) of pseudohypoparathyroidism cases.

### References

- Baheti B, Innani S, Gajre S, et al (2020) Eff-UNet: A novel architecture for semantic segmentation in unstructured environment. In: 2020 IEEE/CVF Conference on Computer Vision and Pattern Recognition Workshops (CVPRW), pp 1473–1481
- Bradski G (2000) The OpenCV Library. *Dr Dobb's Journal of Software Tools*
- Buslaev A, Iglovikov VI, Khvedchenya E, et al (2020) Albumentations: fast and flexible image augmentations. *Information* 11(2):125
- Chen X, Girshick R, He K, et al (2019) Tensormask: A foundation for dense object segmentation. In: *Proceedings of the IEEE/CVF international conference on computer vision*, pp 2061–2069
- Cicero M, Bilbily A (2017) Machine learning and the future of radiology: How we won the 2017 rsna ml challenge. Accessed from https://www16bitai.com/blog/ml-and-future-of-radiology
- Deng J, Dong W, Socher R, et al (2009) Imagenet: A large-scale hierarchical image database. In: *2009 IEEE conference on computer vision and pattern recognition*, Ieee, pp 248–255
- Falcon W, et al (2019) Pytorch lightning. GitHub Note: https://github.com/PyTorchLightning/pytorch-lightning 3(6)
- Greulich WW, Pyle SI (1959) *Radiographic Atlas of Skeletal Development of the Hand and Wrist*. Stanford University Press
- He K, Zhang X, Ren S, et al (2015) Delving deep into rectifiers: Surpassing human-level performance on imagenet classification. In: *Proceedings of the*

IEEE international conference on computer vision, pp 1026–1034

Henschel L, Conjeti S, Estrada S, et al (2020) Fastsurfer-a fast and accurate deep learning based neuroimaging pipeline. *NeuroImage* 219:117,012

Hustinx A, Hellmann F, Sümer et al (2023) Improving deep facial phenotyping for ultra-rare disorder verification using model ensembles. In: 2023 IEEE/CVF Winter Conference on Applications of Computer Vision (WACV), pp 5007–5017, https://doi.org/10.1109/WACV56688.2023.00499

Kingma DP, Ba J (2014) Adam: A method for stochastic optimization. *arXiv preprint arXiv:1412.6980*

Martin DD, Deusch D, Schweizer R, et al (2009) Clinical application of automated greulich-pyle bone age determination in children with short stature. *Pediatric radiology* 39(6):598–607

Pan I, Thodberg HH, Halabi SS, et al (2019) Improving automated pediatric bone age estimation using ensembles of models from the 2017 RSNA machine learning challenge. *Radiology: Artificial Intelligence* 1(6):e190,053. https://doi.org/10.1148/ryai.2019190053, URL https://doi.org/10.1148/ryai.2019190053

Paszke A, Gross S, Massa F, et al (2019) Pytorch: An imperative style, high-performance deep learning library. In: *Advances in Neural Information Processing Systems* 32. Curran Associates, Inc., p 8024–8035

Pizer SM, Amburn EP, Austin JD, et al (1987) Adaptive histogram equalization and its variations. *Computer vision, graphics, and image processing* 39(3):355–368

Pontikos N, Woof W, Veturi A, et al (2022) Eye2gene: prediction of causal inherited retinal disease gene from multimodal imaging using deep-learning https://doi.org/10.21203/rs.3.rs-2110140/v1, URL https://doi.org/10.21203/rs.3.rs-2110140/v1

Tanner JM, Healy MJR, Goldstein H, et al (2001) Assessment of skeletal maturity and prediction of adult height (TW3 method), 3rd edn. W.B. Saunders, London

Wu Y, Kirillov A, Massa F, et al (2019) Detectron2. https://github.com/facebookresearch/detectron2

Yune S, Lee H, Kim M, et al (2019) Beyond human perception: sexual dimorphism in hand and wrist radiographs is discernible by a deep learning model. *Journal of Digital Imaging* 32(4):665–671
